# Unequal effects of health behaviors in adolescence on adult cardiovascular disease and hypertension by family financial situation in the US: A cohort study

**DOI:** 10.64898/2026.03.11.26348128

**Authors:** Nazihah Noor, Josephine Jackisch, Arnaud Chiolero, Kathleen Mullan Harris, Cristian Carmeli

**Affiliations:** Population Health Laboratory (#PopHealthLab), University of Fribourg, Fribourg, Switzerland; Swiss School of Public Health (SSPH+), Zurich, Switzerland; Max Planck Institute for Demographic Research, Rostock, Germany; School of Population and Global Health, McGill University, Montreal, Canada; Department of Sociology and Carolina Population Center, University of North Carolina at Chapel Hill, Chapel Hill, North Carolina, United States of America

## Abstract

**Background:** Adolescent health risk behaviors may have unequal effects on adult cardiovascular diseases (CVDs) and hypertension, with individuals from disadvantaged socioeconomic backgrounds potentially experiencing stronger harmful effects due to heightened susceptibility. Using data from the US National Longitudinal Study of Adolescent to Adult Health, we quantified differential effects of adolescent health behaviors on CVDs and hypertension by family financial situation.

**Methods:** We identified behavior clusters via latent class modeling and estimated effects using double-robust inverse probability weighted methods. Exposures were adolescent health risk behaviors (breakfast skipping, cigarette smoking, frequent alcohol use, and infrequent physical activity), measured at ages 12–19 (1994–1995, n=4452), with parental difficulty paying bills as the effect modifier. CVDs and hypertension were assessed at ages 33–43 (2016–2018) using biomarkers and self-reports.

**Results:** After 21 years, 33.2% of participants had CVD or hypertension. Frequent alcohol use and concurrent cigarette smoking led to 11.1 percentage points more cases (95% CI: 0.1–22.1) among adolescents with parental financial difficulty compared to peers without. Alcohol use alone exhibited a differential effect of 9.7 percentage points (95% CI: 0.8–18.5). The differential effect of breakfast skipping was inconsistent across sensitivity analyses, and no strong evidence was found for infrequent physical activity.

**Conclusions:** These findings from a US cohort indicate that frequent alcohol use in adolescence, particularly with concurrent smoking, increases risk of adult CVD and hypertension unequally based on socioeconomic background. This differential susceptibility suggests that population-level interventions targeting alcohol use and concurrent smoking could help reduce cardiovascular inequalities.

## Introduction

Cardiovascular diseases (CVDs) and hypertension are leading causes of disability and death worldwide (1). These conditions display a social gradient, with individuals from socioeconomically disadvantaged situations facing higher risks of disease and related mortality compared to those from more advantaged socioeconomic circumstances. This disparity is partly attributed to a higher prevalence of health risk behaviors including alcohol consumption, cigarette smoking, poor dietary habits, and physical inactivity among those socially disadvantaged (2).

While the differential exposure to health risk behaviors has been extensively documented, the complementary mechanism of differential vulnerability or susceptibility remains underexplored (3). Differential susceptibility refers to the phenomenon whereby individuals from disadvantaged socioeconomic situations may experience stronger harmful effects of health risk behaviors, contributing to the observed socioeconomic gradient in CVDs and hypertension. Differential susceptibility can arise because of biological embedding of early life adverse experiences (4, 5) or because of the clustering of health risk behaviors during adolescence which then form a lifestyle that persists into adulthood (6, 7). Individuals who experienced material or psychosocial adversities in early life may have a heightened stress response system (8), making them more vulnerable to the negative impacts of health risk behaviors on their health and development of chronic diseases (5, 9, 10). Parental socioeconomic position in early life is critical to determining these early life conditions and thus contributes to the development of socioeconomic health inequalities in adulthood (11).

Despite its potential importance, most studies on differential susceptibility to health behaviors have primarily focused on adults, with limited evidence available on adolescence, a potentially critical period when many health behaviors are established (12–15). Recently, a Finnish cohort study found that socioeconomic background during adolescence can modify the long-term effects of adolescent fruit and vegetable consumption on adult blood pressure levels, although this effect modification was not observed for other health behaviors (16). These emerging findings suggest the need to further examine whether socioeconomic disadvantage in adolescence can modify the long-term effects of other health behaviors on cardiovascular diseases and hypertension across different settings.

Quantifying differential susceptibility to health risk behaviors during adolescence is essential for developing effective public health strategies. Whole-population interventions that achieve the uniform relative reduction in health risk behaviors across socioeconomic groups could yield greater health improvement among the disadvantaged, thus narrowing inequalities. Recognizing greater susceptibility among socioeconomically disadvantaged adolescents could inform the design of targeted policies and programs that provide additional support to these groups, further mitigating health inequalities.

Given the critical role of adolescent behaviors in shaping health outcomes over the life course, this study aims to quantify the differential effect of health risk behaviors among US adolescents on adult CVDs and hypertension based on their family financial situation. We consider family financial ability, here defined as the ability to pay the bills, because it captures household economic situation related to household expenditure and wealth (17). Unlike traditional socioeconomic indicators such as income or education, assessing financial situation can reveal health-related inequalities across all socioeconomic levels, thereby informing public health and policy strategies that reach individuals who might otherwise be overlooked (18, 19). Additionally, financial situation was included in the 2024 American Heart Association and American College of Cardiology report on key data elements and definitions of determinants of health in cardiology (20). Financial strain has been associated with poorer cardiovascular health and higher cardiovascular risk in adulthood, as shown in a recent systematic review and meta-analysis (21). Thus, it is important to examine family financial situation as an effect modifier especially in the US, where over 37% of households in 2024 reported difficulty paying for basic expenses (22).

By examining the differential effects of adolescent health behaviors by family financial situation, this study seeks to expand the understanding of the mechanisms underlying social inequalities in long term health outcomes.

## Methods

### Study and analytic populations

Our target population is adolescents in the United States. The study population was from the National Longitudinal Study of Adolescent to Adult Health (Add Health), a longitudinal and nationally representative sample of US adolescents (https://addhealth.cpc.unc.edu/). Initial data collection was completed through in-home interviews of both participants and their parents in 1994 and 1995 (Wave I; ages 12–21). Since then, four follow up periods of data collection have been completed: Wave II (1995–1996; ages 13–20), Wave III (2001–2002; ages 18–26), Wave IV (2008–2009; ages 24–32), and Wave V (2016–2018; ages 33–43). Each survey wave implemented a stratified 1-level cluster sampling to ensure that the sample is representative of US schools with respect to region of country, urbanicity, school size, school type, and ethnicity (23). Participation rate at all waves is over 70%. The ensuing dataset comprises rich information on demographic, social, familial, socioeconomic, behavioral and adult health measures, making it well suited for this study (23, 24).

The flow chart in Fig. 1 outlines the selection process of the analytic samples. After excluding Wave I participants that were older than 19 (n=190) or with missing age (n=17), our study population included 20583 adolescents. The sample for the cluster analysis comprised 18400 adolescents and was derived by excluding individuals with missing health behaviors (n=367) and with missing sampling design information at Wave I (n=1771). The sample for the causal analysis comprised 4452 adults and was derived by excluding individuals with missing health behaviors in adolescence (n=367), who were not sampled at Wave V for biomarker collection (n=14869), with missing Wave V sampling design information (n=110) and with missing parental financial situation during adolescence (n=740). We excluded individuals with missing health behaviors and parental financial situation during adolescence because we hypothesized missingness not at random for these covariates (25).

**Figure 1.**
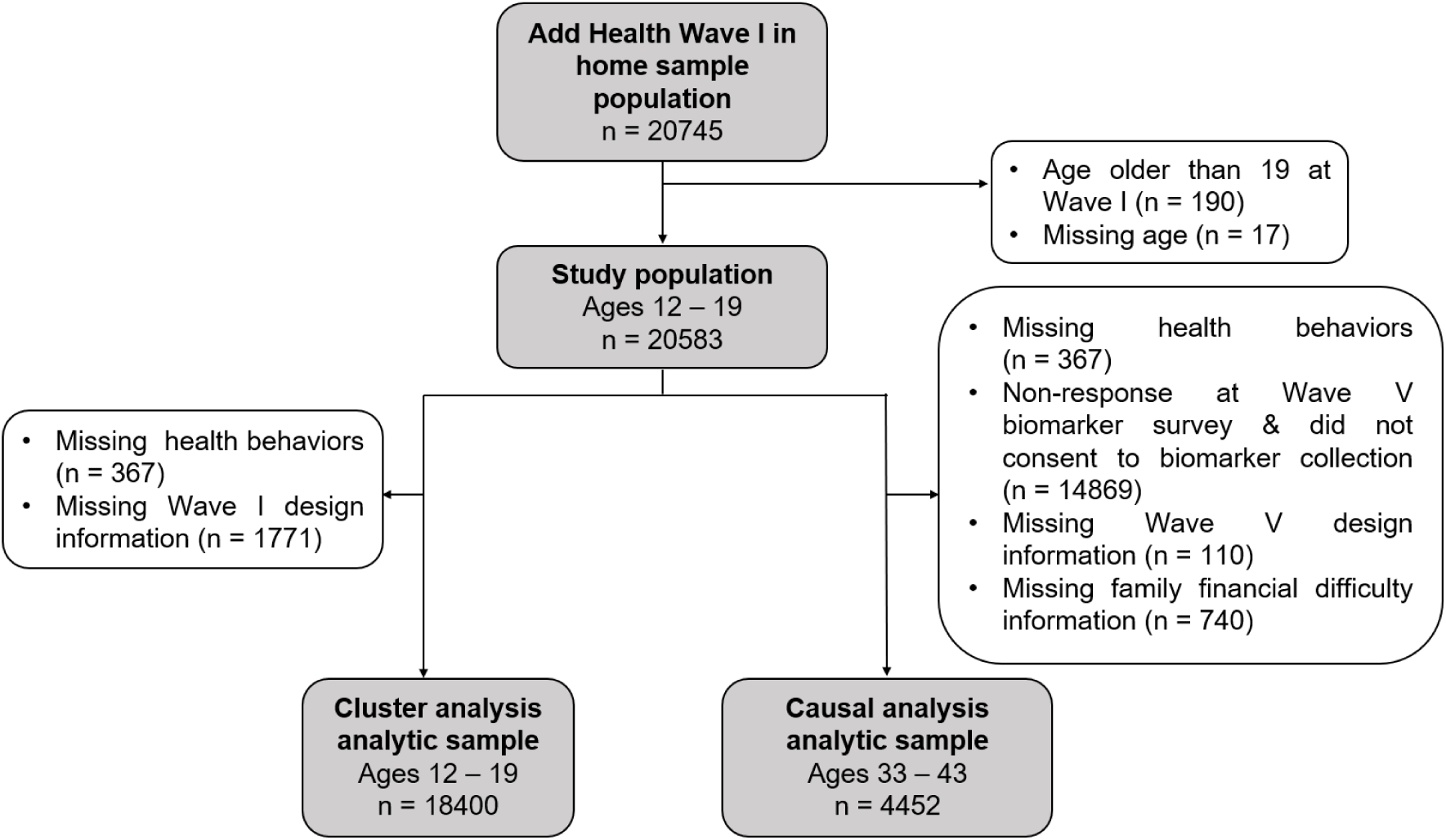
Flow chart of participants selection.

The analytic sample is nationally representative of the US population of adolescents in grades 7–12 in 1994–1995. This is achieved using the Wave V biomarker weights, which adjusts for unequal probabilities of selection in Wave I, attrition across waves, and differential consent for biomarker collection during the Wave V in-home exam (26).

### Causal model and measures

Based on existing literature and the knowledge of the authors (27), we built a conceptual causal model around the main causal path of interest, namely the effect of health behaviors in adolescence (exposure) on CVDs and hypertension in adulthood (outcome), as shown in Figure 2. Potential confounding included socioeconomic factors relative to the family and the neighborhood, parental health behaviors, history of adversities, chronic diseases, mental health disorders and excess weight.

**Figure 2.**
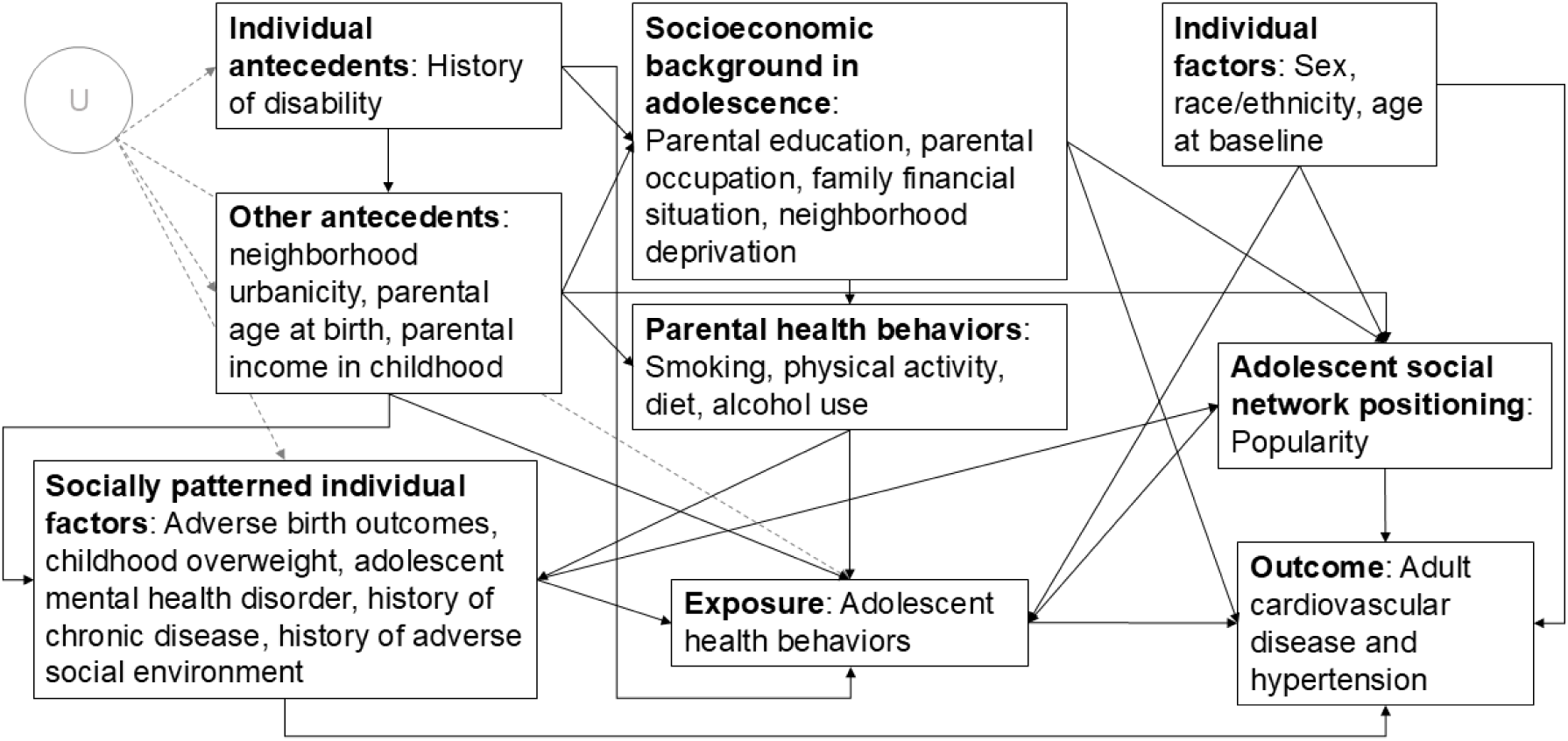
Conceptual causal model for adolescent health behaviors (exposure) to CVDs and hypertension in adulthood (outcome). U stands for other unidentified confounding factors that are not included in the model.

We subsequently established an analytical directed acyclic graph (DAG, see Figure S1 in Supplementary Materials) to identify a subset of confounding factors belonging to the minimally sufficient adjustment set for the causal analysis. The effect modifier of interest, family financial situation, is illustrated within a broader super-node labelled “socioeconomic background in adolescence” in the conceptual causal model shown in Figure 2 and appears as a separate node in the full DAG in Supplementary Figure S1.

#### Exposure: Health behaviors

Health behaviors during adolescence, including alcohol use, cigarette smoking, breakfast eating habits, and physical activity, were assessed using self-reports collected at Wave I (see Table S1 in Supplementary Materials for the specific questions used to measure these exposures).

Physical activity was measured using two indicators: sports frequency and exercise frequency, defined by the number of times respondents engaged in these activities per week. Sports referred to participation in active sports such as baseball, basketball or soccer, whereas exercise captured individual physical activities such as jogging, walking or dancing. Both sports and exercise frequencies were categorized into four levels: never (none), 1–2 times/week (low frequency), 3–4 times/week (moderate frequency), and 5 or more times/week (high frequency). Cigarette smoking was categorized into four levels: never smokers, past smokers, those smoking one cigarette/day, and those smoking multiple cigarettes/day. Alcohol use was categorized into five levels: never drinkers, past drinkers (those who did not drink in the past 12 months but had before), rare drinkers (those who drank alcohol 1–2 days in the past 12 months), moderately frequent drinkers (those who drank once a month or less or 2–3 days a month in the past 12 months), and frequent drinkers (those who drank more often than once a month). Breakfast eating habits were assessed based on whether respondents usually consumed breakfast, categorized into those who had nothing or only coffee/tea, those who had cereal (either alone or with other items), and those who had other types of breakfast foods. For the cluster analysis, all levels of behaviors were used.

For the causal analysis, these behaviors were recoded into risk categories in line with WHO recommendations for low risk health behaviors during adolescence (28) or based on existing evidence on the prevention of chronic diseases (29). Sports and exercise were each assigned to low risk if adolescents practiced frequent physical activity (≥5 times/week), moderate risk if they engaged 3 – 4 times/week, and insufficient/high risk if they participated 0 – 2 times/week. Smoking was designated low risk for never smokers, moderate risk for past smokers, and high risk for current smokers. Alcohol use was deemed low risk for never drinkers, moderate risk for past or rare drinkers, and high risk for moderate or frequent drinkers. Lastly, breakfast habit was dichotomized, where not skipping breakfast was assigned as low risk for those who usually consumed cereal or other foods, and skipping breakfast was assigned as high risk if they had nothing or only coffee or tea.

#### Effect modifier: Family financial situation

Family financial situation was determined using the parent survey in Wave I, where the adolescent’s parent was asked if they had enough money to pay their bills (yes/no). In a sensitivity analysis, we examined equivalized parental gross income (bottom vs top income quintiles).

#### Outcome: CVDs and hypertension

Our outcome is defined as the presence of any of the following conditions: stroke, heart failure, heart disease, atrial fibrillation (collectively grouped as CVDs) and hypertension in adulthood as measured at Wave V, where respondents were aged between 33 and 43 years old. Participants reporting any of these conditions were considered as having CVDs or hypertension. We also conducted a separate analysis where hypertension is the sole outcome.

For hypertension, respondents were flagged as having the condition either through biomarkers, self-reported history of being diagnosed or being on medication. Stroke, heart failure and atrial fibrillation were identified through self-reported previous diagnosis by a healthcare professional. Heart disease was identified through self-reported previous diagnosis by a healthcare professional or history of heart surgery for clogged coronary arteries.

#### Confounding factors

For our causal analysis, the minimally sufficient adjustment set identified using the DAG included age at baseline survey, sex, race, adverse birth outcome, history of chronic disease diagnosis, childhood overweight, adolescent mental disorder, history of adverse social environment, adolescent social network position, neighborhood deprivation, parental education, family financial situation in adolescence and parental smoking.

Age at baseline survey was measured at Wave I via self-reported date of birth and categorized into early adolescence (12–14 years) and mid-late adolescence (15–19 years) to reflect meaningful developmental differences in autonomy, exposure to health behaviors and susceptibility to social influences in different stages of adolescence (30). Sex at birth was self-reported at Wave I and categorized as male or female. Race/ethnicity was measured through self-report and interviewer-identified at Wave I and categorized into (non-Hispanic) White, (non-Hispanic) Black, Hispanic and (non-Hispanic) Other.

Adverse birth outcome was self-reported at Wave V, coded yes for those born preterm (before week 37 of gestational age) and no otherwise. History of chronic disease was self-reported at Wave V, defined as diagnosis of hypertension, diabetes, stroke, cardiac failure, cancer or asthma by a medical professional before the adolescent age at baseline (Wave I). Pre-adolescent overweight was measured as a diagnosis of obesity before the age of 16 by a medical professional, self-reported at Wave V.

Adolescent mental disorder was measured at Wave I as the presence of depressive symptoms or suicidal ideation. Depressive symptoms were measured using the frequency of experiencing three feelings: could not shake off the blues even with help from family and friends, felt depressed during the past 7 days, and felt sad during the past 7 days. Responses were scored as 0 (never or rarely), 1 (sometimes), 2 (a lot of the time), or 3 (most/all of the time). Participants were considered to have depressive symptoms if they scored 2 or higher on any of these three questions. Suicidal ideation was measured by whether the respondent seriously thought about committing suicide in the past 12 months. Respondents were classified as having a mental disorder if they had either depressive symptoms or suicidal ideation.

History of adverse social environment was defined as the presence of any of the following adversities: incarceration of a biological parent before age at baseline self-reported at Wave IV, school expulsion self-reported at Wave I, or physical/sexual abuse self-reported at Wave IV. Adolescent social network position is measured as popularity (31), which is the number of friend nominations received by a respondent from other students in their school, collected at the in-school survey component of Wave I.

Neighborhood deprivation was measured at Wave I using a constructed neighborhood disadvantage score, based on Add Health participants’ addresses when they were first interviewed in 1994–1995. The score was created based on five measures, namely the proportion of female-headed households, individuals receiving public assistance, adults with less than a high school education and adults who were unemployed. The score was z-transformed to have zero mean and unit variance, and then categorized into terciles (20).

Parental education was dichotomized by either at least one parent who had graduated from a college or university or did not, using information from both the parent survey and the adolescent survey in Wave I. Parental smoking was determined using the parent survey in Wave I, based on whether there was a smoker in the household (yes/no).

### Statistical analysis

We implemented the analysis in two parts, first a cluster analysis followed by the causal analysis. All analytical methods used Add Health’s sampling design to account for unequal probability of selection and attrition.

#### Cluster analysis

We carried out a latent class analysis (LCA) to assess the presence of co-occurring health behaviors during adolescence. The LCA provided insights into whether health behaviors in our causal analysis should be analyzed solely as individual exposures or whether any co-occurring behavior patterns should also be considered. In other words, the aim of this analysis was to describe health behavior patterns among adolescents in our study population, not to use the latent classes as exposures in our causal analysis. Our approach is aligned with health lifestyles theory, which conceptualizes health behaviors as co-occurring practices that emerge from the interaction of individual choices and structural conditions which often form meaningful behavioral patterns rather than independent, additive risks (7, 32, 33). We used the gsem package in Stata 18 to perform the LCA and determined the best number of classes by model fit statistics.

#### Causal analysis

We quantified the effect modification of health behaviors on CVDs or hypertension by family financial difficulty. We estimated additive risk differences to quantify the excess burden of CVDs and hypertension across socioeconomic strata as they are commensurable to the prevalence of these diseases in the target population and are then directly interpretable for assessing the impact of potential public health interventions (34, 35). Specifically, our estimand was the difference in conditional average causal effects (cACE) between adolescents without family financial difficulty and adolescents with family financial difficulty. Each cACE represents the marginal difference in potential outcomes between counterfactual scenarios: one in which all adolescents are exposed to high risk health behavior and another in which all adolescents are exposed to low risk health behavior. As a secondary analysis, we also estimated the cACE comparing a counterfactual scenario where all adolescents were exposed to moderate risk health behavior to the counterfactual scenario where all adolescents were exposed to low risk health behavior.

To estimate the cACE, we used a doubly robust method (36). First, we applied a multinomial logistic regression to model the polytomous exposures (sports, exercise, smoking and alcohol use) and binary logistic regression for the dichotomous breakfast skipping variable. We used these models to compute stabilized inverse probability weights, which account for measured confounding and ensure balance across exposure groups (see Figure S2 in the Supplementary Material for assessment of weights distribution and of exposure balance before and after weighting). Second, we performed weighted logistic regression to predict the outcomes with the exposure and family financial situation (and their product term), and the measured confounding factors. Internal validity of the estimates requires various assumptions: no residual confounding, consistency, positivity, no measurement error, and correct specification of either the exposure or the outcome regression models (37). We measured effect magnitudes on the additive scale via risk differences (RD) per 100 persons or percentage for each cACE and their difference (35).

Compatibility intervals (CI) (38) were generated by 1000 bootstrapped sampling designs with the Rao-Wu-Yue-Beaumont method (39). Within each bootstrapped design, the effect estimates were the average of estimates from 100 imputed data sets obtained using multiple imputation by chained equations and hypothesizing missingness at random for neighborhood deprivation, adverse birth outcome, adolescent mental disorder, history of chronic diseases, history of social adversity, parental education, household smoking, childhood overweight, and adolescent social network position (see Table 1 for the numbers missing that were imputed and supplementary material for more details on the imputation model). Causal analyses were conducted in R version 4.4.0.

**Table 1.**
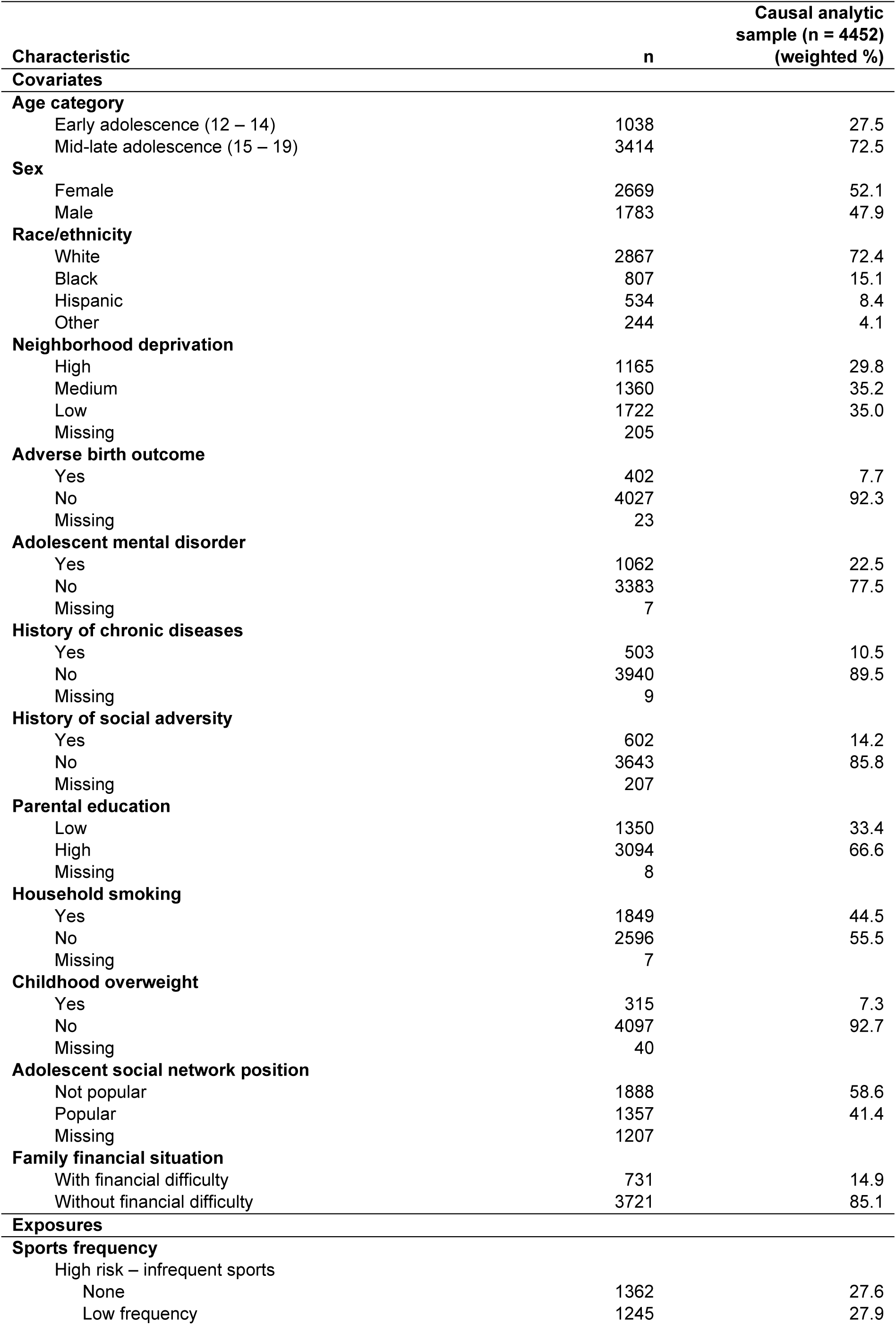
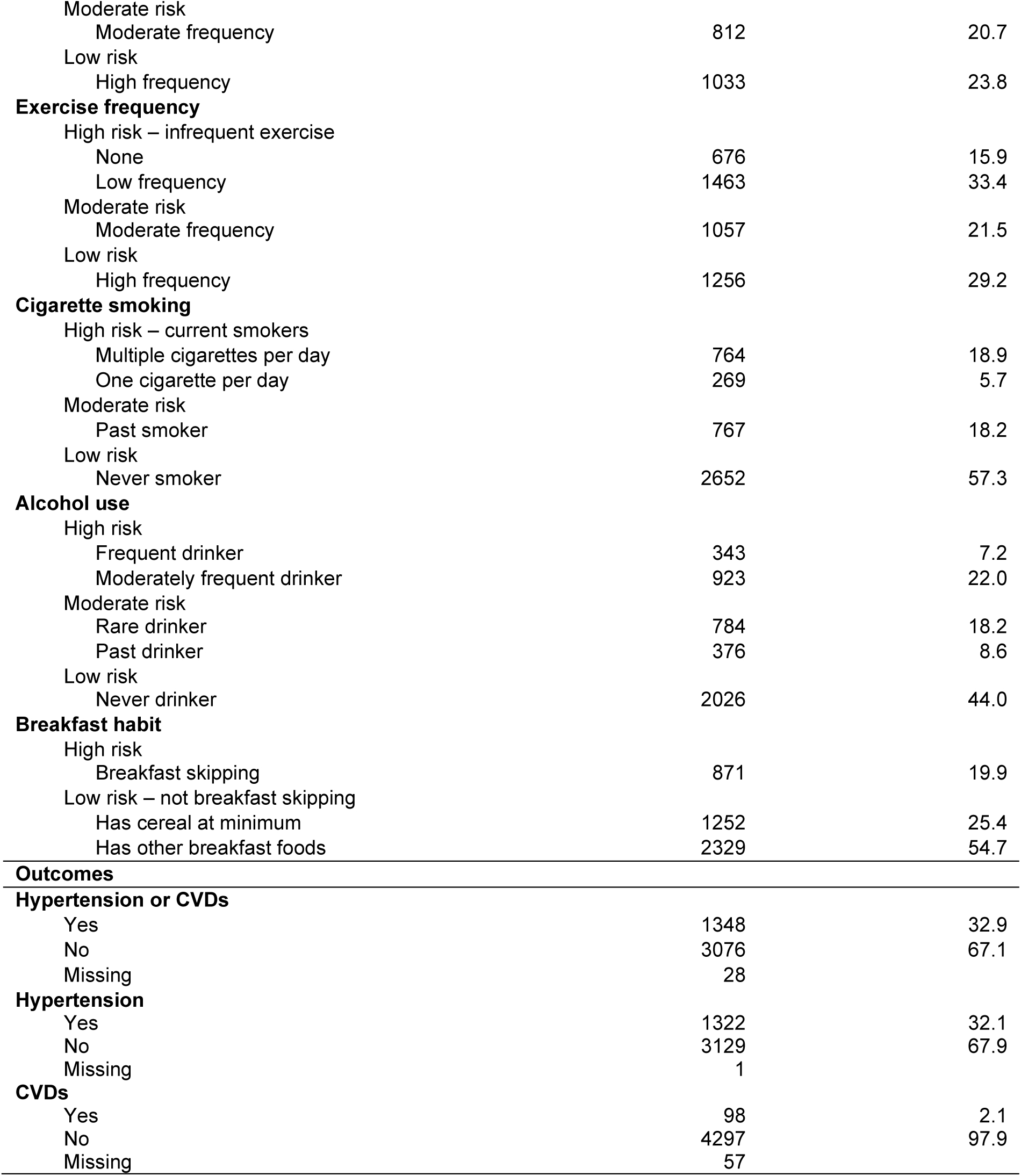
Summary of causal analytic sample characteristics.

| Characteristic | n | Causal analytic sample (n = 4452) (weighted %) |
| --- | --- | --- |
| **Covariates** |  |  |
| **Age category** |  |  |
| Early adolescence (12 – 14) | 1038 | 27.5 |
| Mid-late adolescence (15 – 19) | 3414 | 72.5 |
| **Sex** |  |  |
| Female | 2669 | 52.1 |
| Male | 1783 | 47.9 |
| **Race/ethnicity** |  |  |
| White | 2867 | 72.4 |
| Black | 807 | 15.1 |
| Hispanic | 534 | 8.4 |
| Other | 244 | 4.1 |
| **Neighborhood deprivation** |  |  |
| High | 1165 | 29.8 |
| Medium | 1360 | 35.2 |
| Low | 1722 | 35.0 |
| Missing | 205 |  |
| **Adverse birth outcome** |  |  |
| Yes | 402 | 7.7 |
| No | 4027 | 92.3 |
| Missing | 23 |  |
| **Adolescent mental disorder** |  |  |
| Yes | 1062 | 22.5 |
| No | 3383 | 77.5 |
| Missing | 7 |  |
| **History of chronic diseases** |  |  |
| Yes | 503 | 10.5 |
| No | 3940 | 89.5 |
| Missing | 9 |  |
| **History of social adversity** |  |  |
| Yes | 602 | 14.2 |
| No | 3643 | 85.8 |
| Missing | 207 |  |
| **Parental education** |  |  |
| Low | 1350 | 33.4 |
| High | 3094 | 66.6 |
| Missing | 8 |  |
| **Household smoking** |  |  |
| Yes | 1849 | 44.5 |
| No | 2596 | 55.5 |
| Missing | 7 |  |
| **Childhood overweight** |  |  |
| Yes | 315 | 7.3 |
| No | 4097 | 92.7 |
| Missing | 40 |  |
| **Adolescent social network position** |  |  |
| Not popular | 1888 | 58.6 |
| Popular | 1357 | 41.4 |
| Missing | 1207 |  |
| **Family financial situation** |  |  |
| With financial difficulty | 731 | 14.9 |
| Without financial difficulty | 3721 | 85.1 |
| **Exposures** |  |  |
| **Sports frequency** |  |  |
| High risk – infrequent sports |  |  |
| None | 1362 | 27.6 |
| Low frequency | 1245 | 27.9 |
| Moderate risk |  |  |
| Moderate frequency | 812 | 20.7 |
| Low risk |  |  |
| High frequency | 1033 | 23.8 |
| **Exercise frequency** |  |  |
| High risk – infrequent exercise |  |  |
| None | 676 | 15.9 |
| Low frequency | 1463 | 33.4 |
| Moderate risk |  |  |
| Moderate frequency | 1057 | 21.5 |
| Low risk |  |  |
| High frequency | 1256 | 29.2 |
| **Cigarette smoking** |  |  |
| High risk – current smokers |  |  |
| Multiple cigarettes per day | 764 | 18.9 |
| One cigarette per day | 269 | 5.7 |
| Moderate risk |  |  |
| Past smoker | 767 | 18.2 |
| Low risk |  |  |
| Never smoker | 2652 | 57.3 |
| **Alcohol use** |  |  |
| High risk |  |  |
| Frequent drinker | 343 | 7.2 |
| Moderately frequent drinker | 923 | 22.0 |
| Moderate risk |  |  |
| Rare drinker | 784 | 18.2 |
| Past drinker | 376 | 8.6 |
| Low risk |  |  |
| Never drinker | 2026 | 44.0 |
| **Breakfast habit** |  |  |
| High risk |  |  |
| Breakfast skipping | 871 | 19.9 |
| Low risk – not breakfast skipping |  |  |
| Has cereal at minimum | 1252 | 25.4 |
| Has other breakfast foods | 2329 | 54.7 |
| **Outcomes** |  |  |
| **Hypertension or CVDs** |  |  |
| Yes | 1348 | 32.9 |
| No | 3076 | 67.1 |
| Missing | 28 |  |
| **Hypertension** |  |  |
| Yes | 1322 | 32.1 |
| No | 3129 | 67.9 |
| Missing | 1 |  |
| **CVDs** |  |  |
| Yes | 98 | 2.1 |
| No | 4297 | 97.9 |
| Missing | 57 |  |

#### Sensitivity analyses

To assess sensitivity of findings when using a different operationalization of the effect modifier, we ran the analysis when financial situation was measured through levels of equivalized parental gross income (bottom income quintile vs top income quintile). Additionally, to assess sensitivity of our causal estimates to unmeasured confounding, we estimated differential effects on a negative control outcome (40), that we identified in the self-reported abstention from voting in political elections.

## Results

### Characteristics of the analytic sample

Table 1 summarizes the characteristics of adolescents in our causal analytic sample, with weighted percentages provided in parentheses. At baseline, 72.5% of adolescents were aged 15‒19 and 52.1% were female. The racial/ethnic composition was predominantly White (72.4%), followed by Black (15.1%), Hispanic (8.4%) and other (4.1%). In terms of early life health conditions, 7.3% had childhood obesity, 10.5% had a history of chronic disease, and 22.5% had an adolescent mental disorder. Most adolescents were from socioeconomically advantaged backgrounds, with 66.6% of them from highly educated parents and 85.1% from families without financial difficulty. Approximately half of adolescents had high risk levels of physical activity (55.5% no sports or low frequency and 49.3% no exercise or low frequency). Almost a quarter of adolescents reported to be current smokers (24.6%). 29.2% of adolescents engaged in high risk alcohol use. Almost one in five adolescents regularly skips breakfast (19.9%). After an average follow-up of 21 years in early midlife, the prevalence of CVDs and hypertension among the adults aged 33‒43 was 32.9%.

Infrequent sports and breakfast skipping were more common among adolescents from families with financial difficulty (66.2% and 23.3%, respectively) compared to their advantaged peers (57.1% and 18.8%, respectively), while smoking and alcohol use showed minimal differences between the groups (Supplementary Table S2). Finally, the prevalence of CVDs and hypertension was higher among adolescents from families with financial difficulty compared to their advantaged peers (36.6% and 29.7%, respectively).

### Clustering of health behaviors

The LCA identified two classes of health lifestyles among the Wave I Add Health adolescents (see Supplementary Material). Class 1 represents 60% of adolescents, characterized by high probabilities of engaging in healthier behavior, specifically never drinking alcohol (0.69), never smoking (0.82), and having breakfast usually (0.86). Class 2 represents 40% of adolescents, who exhibit low probability of engaging in healthier behaviors, particularly for never and past smoking/alcohol use (0.40 and 0.13 respectively).

Based on the LCA, alcohol use and cigarette smoking appear to co-occur, whereby adolescents who have frequent or moderately frequent alcohol use are also more likely to be current cigarette smokers and vice versa for never or past smoking/alcohol. Similar patterns of behavior clustering were observed with a three-class model and a four-class model (Supplementary Material). Guided by the results of the LCA, we analyzed the effect of concurrent smoking and alcohol in addition to the single health risk behaviors in our causal analysis. Exposure to concurrent smoking and alcohol use was operationalized as high risk for being a current smoker and a frequent or moderately frequent alcohol user (n=648, weighted % = 15.9%), low risk for being a never smoker and never drinker (n=1707, weighted % = 38.2%), and moderate risk otherwise (n=2097, weighted % = 45.9%).

### Differential effects of health risk behaviors on CVDs and hypertension

Estimated average causal effects of high risk health behaviors among adolescents with and without family financial difficulty are displayed in Figure 3 and Supplementary Table S5. Among adolescents from advantaged families, the effects of infrequent sport, infrequent exercise, current smoking, moderate or frequent alcohol use, breakfast skipping, and the combination of smoking and alcohol use were small or negligible (under 5%). In contrast, among adolescents from disadvantaged families, the effects of breakfast skipping, alcohol use, and concurrent smoking and alcohol use were substantial.

**Figure 3.**
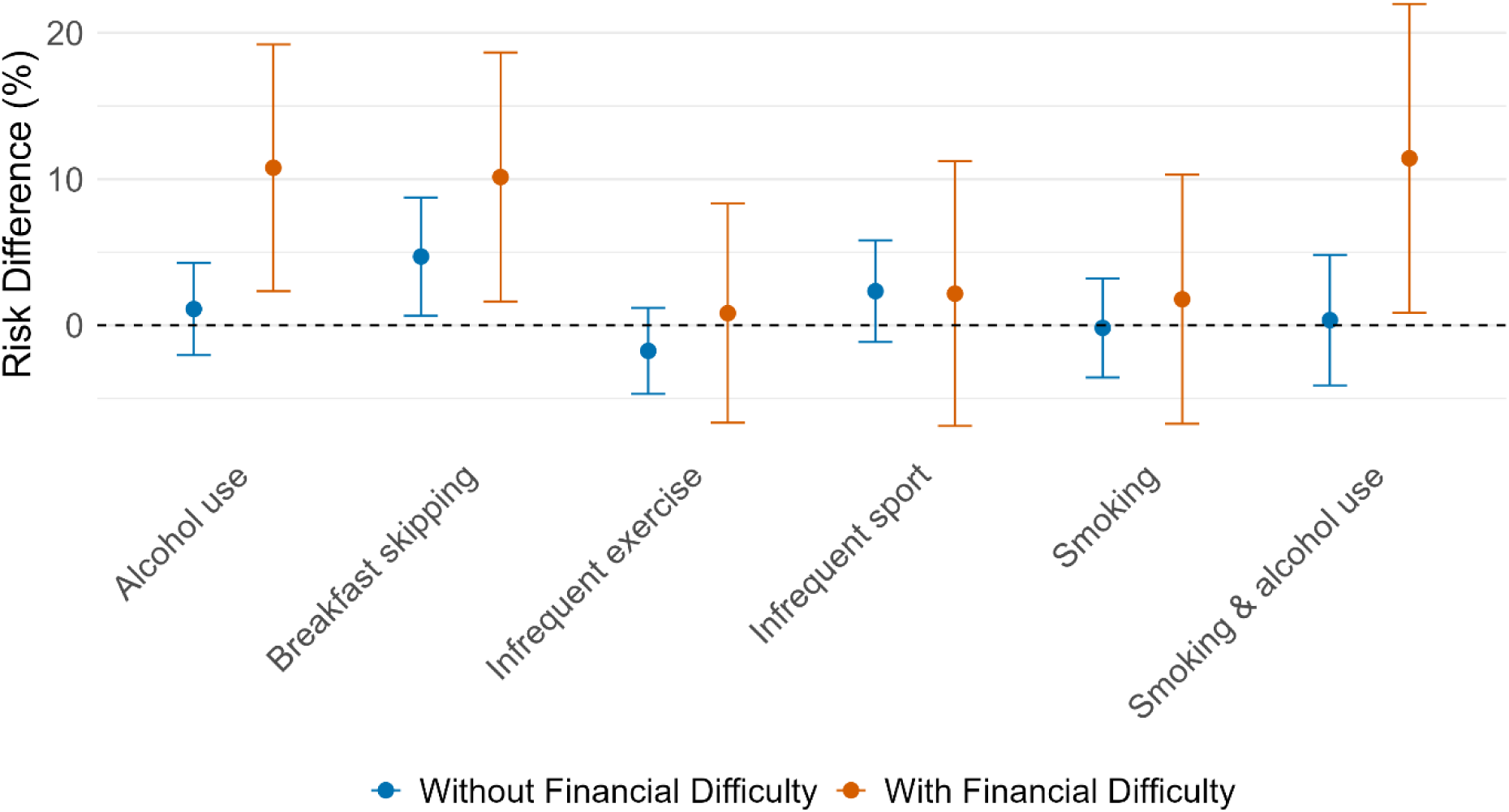
Average causal effect of high risk health behaviors on CVDs or hypertension among adolescents with family financial difficulty and among adolescents without family financial difficulty. Magnitude is measured via risk difference per 100 persons (% percentage points). Alcohol use encompasses frequent and moderately frequent drinking. Infrequent exercise/sports encompass low frequency and no exercise/sports. Smoking encompasses current smoking (multiple cigarettes or one cigarette per day).

Specifically, breakfast skipping resulted in an additional 10.1 percentage points (95% CI: 1.6–18.7) CVD or hypertension cases for those with parental financial difficulty, compared to an additional 4.7 percentage points (95% CI: 0.7–8.7) among those without parental financial difficulty. Moderate or frequent alcohol use led to 10.8 percentage points (95% CI: 2.3–19.2) more CVD or hypertension cases among disadvantaged adolescents compared to 1.1 percentage points (95% CI: -2.0–4.3) among advantaged adolescents. The effect of concurrent smoking and alcohol use resulted in 11.4 percentage points (95% CI: 0.9–22.0) among those with parental financial difficulty, whereas it was only 0.4 percentage points (95% CI: -4.1–4.8) among adolescents without parental financial difficulty.

The effect modification due to family financial difficulty is displayed in Figure 4. Breakfast skipping led to an excess of 5.4 percentage points (95% CI: -3.7–14.6) CVD or hypertension cases for adolescents with financial difficulties compared to those without. Moderate or frequent alcohol use had an excess of 9.7 percentage points (95% CI: 0.8–18.5), while concurrent smoking and alcohol use had an excess of 11.1 percentage points (95% CI: 0.1–22.1). Other high risk health behaviors had negligible effect modification, indicating that their effect on CVDs or hypertension was not substantially different between socioeconomically advantaged and disadvantaged adolescents.

**Figure 4.**
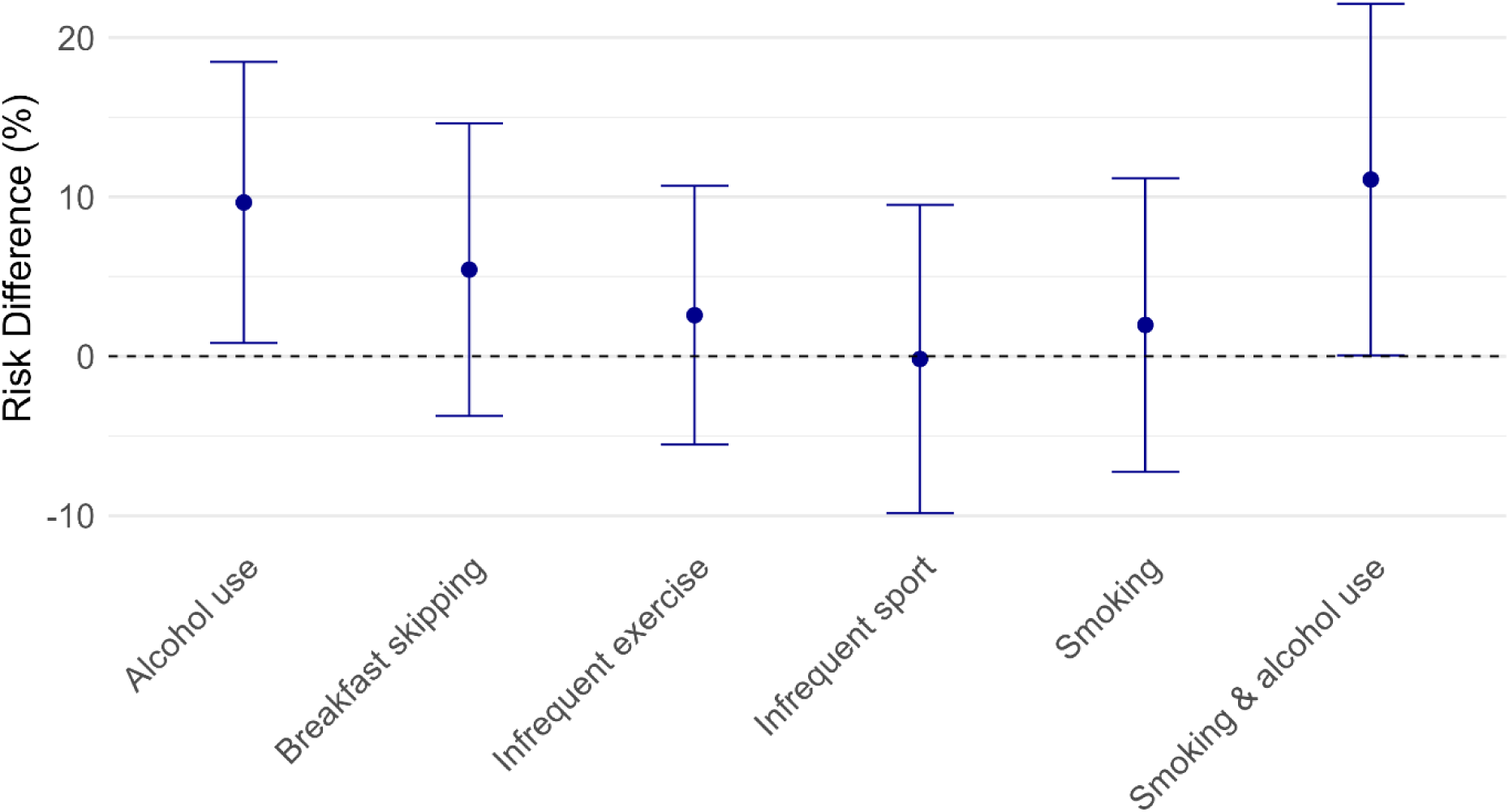
Effect modification associated with family financial situation. The effect modification is the difference in average causal effect between those without family financial difficulty compared to those with family financial difficulty. Alcohol use encompasses frequent and moderately frequent drinking. Infrequent exercise/sports encompass low frequency and no exercise/sports. Smoking encompasses current smoking (multiple cigarettes or one cigarette per day).

The analysis comparing moderate risk to low risk adolescent health behaviors showed variable patterns across exposures (Supplementary Table S6). While moderately frequent exercise and past smoking resulted in effects with small magnitudes similarly to what observed for the corresponding high risk behaviors, moderately frequent sport among adolescents from families with financial difficulty experiencing resulted in a larger effect (11.8 percentage points, 95% CI: -0.6–24.3) compared to that among their advantaged peers (2.3 percentage points, 95% CI: -2.5–7.1). Rare or past alcohol use resulted in a smaller effect among disadvantaged adolescents compared to that observed for moderate or frequent alcohol use, leading to a reduced magnitude of effect modification (5.2 percentage points, 95% CI: -3.4–13.7). A similar pattern was observed for the joint effect of smoking and alcohol use, with an excess risk difference of 8.5 percentage points (95% CI: 0.1–17.0) for socioeconomically disadvantaged adolescents. Given the heterogeneity in smoking and alcohol use patterns within the moderate risk group, we omit this contrast.

When considering hypertension as the sole outcome, the patterns of effect modification were consistent with the main analysis (Supplementary Table S7).

### Sensitivity analyses

Substituting family financial difficulty with equivalized gross parental income as the effect modifier reaffirmed the results of our main analysis for alcohol use and concurrent smoking and alcohol use (Supplementary Figures S4 and S5). The differential effect of alcohol use was 10.8 percentage points (95% CI: 2.3–19.2), which is consistent with the main analysis. Notably, there was a larger effect modification size for concurrent smoking and alcohol use compared to the main analysis, which rose to 27.3 percentage points (95% CI: 12.2–42.4) when comparing the bottom income quintile group to the top income quintile group. Conversely, the effect modification for breakfast skipping became -2.4 percentage points (95% CI: -15.4–10.6), indicating variability in this estimate.

Additionally, as displayed in Supplementary Table S4, the differential effect of alcohol use during adolescence on a negative control outcome (adult abstention from voting) was -0.8 percentage points (95% CI: -8.2–6.7), indicating that the main findings cannot be explained by unmeasured confounding. Similarly, the differential effect of concurrent smoking and alcohol use during adolescence on adult abstention from voting was 0.5 percentage points (95% CI: -10.3–11.4), similarly indicating that the main findings are unlikely due to unmeasured confounding. In contrast, breakfast skipping showed a larger differential effect on the negative control outcome (9.1 percentage points, 95% CI: 0.6–17.6 among disadvantaged adolescents), which is similar to the magnitude observed in the main analysis. Thus, these results suggest that findings related to breakfast skipping could be attributable to unmeasured confounding.

## Discussion

In a nationally representative sample of US adolescents, frequent alcohol consumption and concurrent cigarette smoking increased the 20-year risk of CVDs or hypertension more among adolescents from families with financial difficulty than their advantaged peers. Additionally, breakfast skipping was observed to have an unequal effect on CVDs and hypertension. However, sensitivity analysis using a negative control outcome suggested that this differential effect of breakfast skipping may be explained by unmeasured confounding factors. There was no strong evidence of differential effects of adolescent smoking and of infrequent physical activity by family financial situations.

Our main finding on the unequal effects of adolescent lifestyle pattern composed of concurrent smoking and alcohol use aligns with and expand upon existing literature. Hitherto, the evidence on differential effects of smoking and alcohol use on CVDs and hypertension is largely limited to adults. A meta-analysis of three studies reported that engaging in multiple high risk health behaviors (at least three of the following: poor diet, physical inactivity, smoking, alcohol consumption, short sleep duration, sedentary time and television viewing time) increases the risk of cardiovascular mortality and incidence more in adults with socioeconomic disadvantage than those with socioeconomic advantage (41). Our study provides novel evidence of differential susceptibility to multiple health risk behaviors by socioeconomic situation along two dimensions: first, by identifying the specific high risk health behaviors of concurrent frequent alcohol consumption and cigarette smoking, and second, by providing evidence on adolescents. Given that concurrent smoking and alcohol use form a distinct lifestyle pattern, interventions should target both behaviors together rather than separately, particularly given the unequal effects of this pattern on cardiovascular health. Public health interventions must also consider the underlying sociocultural drivers that shape concurrent smoking and alcohol use, such as peer influences and structural factors (42, 43).

The unequal effect of alcohol use on CVDs and hypertension for adolescents from disadvantaged backgrounds also aligns with previous discussions on the alcohol harm paradox, whereby disadvantaged groups experience disproportionate harm despite similar or lower alcohol consumption (13). Previous studies have demonstrated differential effects in alcohol-related cardiovascular mortality, with disadvantaged groups facing greater risks despite comparable consumption levels (13, 44, 45). While these studies examined alcohol use in adulthood and its impact primarily on CVD mortality, our findings contribute new evidence by showing that socioeconomic disparities in susceptibility to alcohol use emerge as early as adolescence and extend to long-term CVD and hypertension development. However, our findings differ from those of a recent Finnish cohort study which found little evidence of differential effects of adolescent alcohol use on adult cardiometabolic health (16). This may be partly explained by differences in the operationalization of alcohol use, where in our study, we distinguish between never drinkers and past drinkers, an important distinction given evidence that former drinkers have higher cardiovascular risk than lifetime abstainers (46). In addition, the Finnish study examined parental education and occupation as effect modifiers, whereas the present study utilizes family financial difficulty, which may reflect more immediate material hardship. Differences in alcohol consumption patterns and health systems between Finland and the US may further contribute to the diverging observations.

When cigarette smoking was considered separately, we did not find robust evidence for differential effects. Findings on differential effects of smoking on CVDs and hypertension from previous studies are also mixed, with no clear evidence for a differential effect of smoking across socioeconomic groups (12, 47, 48).

A meta-analysis of five cohort studies reported an increased risk of CVDs associated with skipping breakfast among adults (49), supporting previous findings that (49) skipping breakfast has harmful effects for CVDs and hypertension (29). However, our findings were not consistent across sensitivity analyses, particularly when using a negative control outcome. One meta-analysis of two randomized controlled trials reported no effect of breakfast skipping on hypertension among adults (50). Additionally, a longitudinal observational study among UK adolescents aged 11 – 13 reported that after 10 years of follow-up, systolic blood pressure levels were similar between those who skipped breakfast compared to those who had breakfast regularly (51). Overall, these findings, along with our results provide limited or insufficient evidence of an effect of adolescent breakfast skipping on adult CVDs and hypertension and of its differential effect.

Similarly, infrequent physical activity, measured as either low frequency to no sport or exercise among US adolescents, did not show strong and consistent effects on adult CVDs and hypertension. This may point to either a downward bias related to social desirability-related overreporting of frequent physical activity among those who are less frequently active (51) or the crudeness of questionnaire-based assessment of physical activity (52). Future studies with more objective and thorough measurements of physical activity, for example based on accelerometer, are warranted.

Additionally, the role of differential health behavior trajectories from adolescence to adulthood is critical in understanding how cardiovascular and hypertensive disparities emerge. Adolescents from disadvantaged backgrounds are more likely to continue risky health behaviors, such as being physically inactive, smoking and drinking excessively into adulthood, while their socioeconomically advantaged peers are more likely to discontinue these behaviors as they transition into young adulthood (53, 54). These differing behavior trajectories could provide additional insights into the heightened susceptibility to joint smoking and alcohol use observed among socioeconomically disadvantaged adolescents. Future research is needed to examine this possibility.

Our study adds further understanding regarding the mechanisms underlying socioeconomic inequalities in CVDs and hypertension. Previous studies have consistently demonstrated the relevance of differential exposure to health behaviors, most notably smoking, among adults (2). Our findings indicate that differential susceptibility to frequent alcohol use and concurrent cigarette smoking among US adolescents, contributed to socioeconomic disparities in long-term cardiovascular diseases and hypertension. This highlights that health outcomes could be socially graded without social gradients in the prevalence of health behaviors, emphasizing the relevance of examining both mechanisms (5). While these mechanisms are complementary, they offer distinct policy entry points to tackle health inequalities. In the presence of differential exposure, targeted population health promotion equalizing smoking and alcohol use would mitigate inequalities. In contrast, where differential susceptibility is present, whole-population interventions that achieve the same relative reduction of smoking and alcohol consumption across socioeconomic groups would yield greater health improvement among the disadvantaged, thus narrowing inequalities.

Our study has some limitations that should be considered. First, while we explicitly and carefully identified all potential confounding factors to the best of our knowledge to establish a causal relationship, there may still be unmeasured confounding that could result in biasing our effect estimates. Additionally, some residual confounding bias could also stem from the crude operationalization of some of the measured confounding factors. However, findings from a negative control outcome indicated that the differential effect of joint smoking and alcohol use is unlikely to be explained by residual confounding. Second, as mentioned for physical activity, reliance on self-reported data for socially sensitive behaviors as smoking and alcohol consumption may introduce reporting errors, potentially biasing upward or downward the estimated differential effect on CVDs and hypertension. Relatedly, some early life confounding factors including adolescent obesity and adverse birth outcomes were measured retrospectively in later waves, which may result in some recall bias. Third, participants without biomarker assessments at Wave V were excluded because measured blood pressure was required to define the hypertension outcome. These biomarker data were missing due to the sampling design rather than participant characteristics, so the exclusion is unlikely to bias our estimates but may reduce statistical precision. Fourth, there could be some bias from the misspecification of either the exposure or the outcome parametric models. Finally, the consistency assumption necessary for the validity of the cACE estimation might be violated as we are not measuring the effect of a specific intervention but rather a potential weighted mean effect of interventions that would change a health behavior.

Strengths of our study include the use of a nationally representative sample of US adolescents enhancing the generalizability of our findings, and the prospective design with long follow-up period of 20 years from adolescence to mid-life. Additionally, the integration of latent class analysis with contemporary methods for causal inference from observational data provides a robust approach to understanding the complex interplay between socioeconomic factors and health behaviors, offering a more comprehensive perspective on their relationship with long-term health outcomes.

This study has two main implications for CVD and hypertension prevention. Our findings indicate that frequent alcohol use and concurrent cigarette smoking during adolescence have a more severe impact on the occurrence of cardiovascular diseases and hypertension for those from socioeconomically disadvantaged families compared to those from advantaged backgrounds. If confirmed by further research, this unequal effect of high risk behaviors points to the potential for whole-population interventions targeting alcohol use and the combination of concurrent smoking and alcohol use to mitigate inequalities in cardiovascular and hypertensive outcomes. Furthermore, acknowledging a greater susceptibility to high risk health behaviors among socioeconomically disadvantaged adolescents could guide the development of targeted policies and programs that provide additional support to these groups to address and mitigate their heightened susceptibility.

## Data Availability

The Add Health data are available in public‑use and restricted‑use formats. Restricted‑use data, including the full cohort, require an approved contract. Information on accessing Add Health data is available at https://addhealth.cpc.unc.edu/data/.

## Acknowledgements

This research uses data from Add Health, a program project directed by Kathleen Mullan Harris and designed by J. Richard Udry, Peter S. Bearman, and Kathleen Mullan Harris at the University of North Carolina at Chapel Hill, and funded by grant P01-HD31921 (Harris) from the Eunice Kennedy Shriver National Institute of Child Health and Human Development, with cooperative funding from 23 other federal agencies and foundations. No direct support was received from grant P01-HD31921 for this analysis.

## Supplementary Material

1. Directed Acyclic Graph (Supplementary Figure S1)
2. Add Health Questions used for Health Behaviors (Supplementary Table S1)
3. Standardized Mean Differences (Supplementary Figure S2)
4. Inverse Probability Weights and Multiple Imputation Models
5. Inequalities in Exposures and Outcomes (Supplementary Table S2)
6. Latent Class Analyses (Supplementary Table S3 and Figure S3)
7. Sensitivity Analyses (Supplementary Figure S4 and S5, Table S4)
8. Main Results Tables (Supplementary Tables S5, S6, S7)

## Non-standard Abbreviations and Acronyms

cACE: Conditional average causal effect
CI: Compatibility interval
CVDs: Cardiovascular diseases
DAG: Directed acyclic graph
LCA: Latent class analysis
RD: Risk difference
UK: United Kingdom
US: United States
WHO: World Health Organization

