## Supplementary Material for "Unequal effects of health behaviors in adolescence on adult cardiovascular disease and hypertension by family financial situation in the US: A cohort study"

Postal address: #PopHealthLab, University of Fribourg, 41 Route des Arsenaux, 1700 Fribourg, Switzerland

#### Supplementary Material

### 1. Directed Acyclic Graph (DAG)

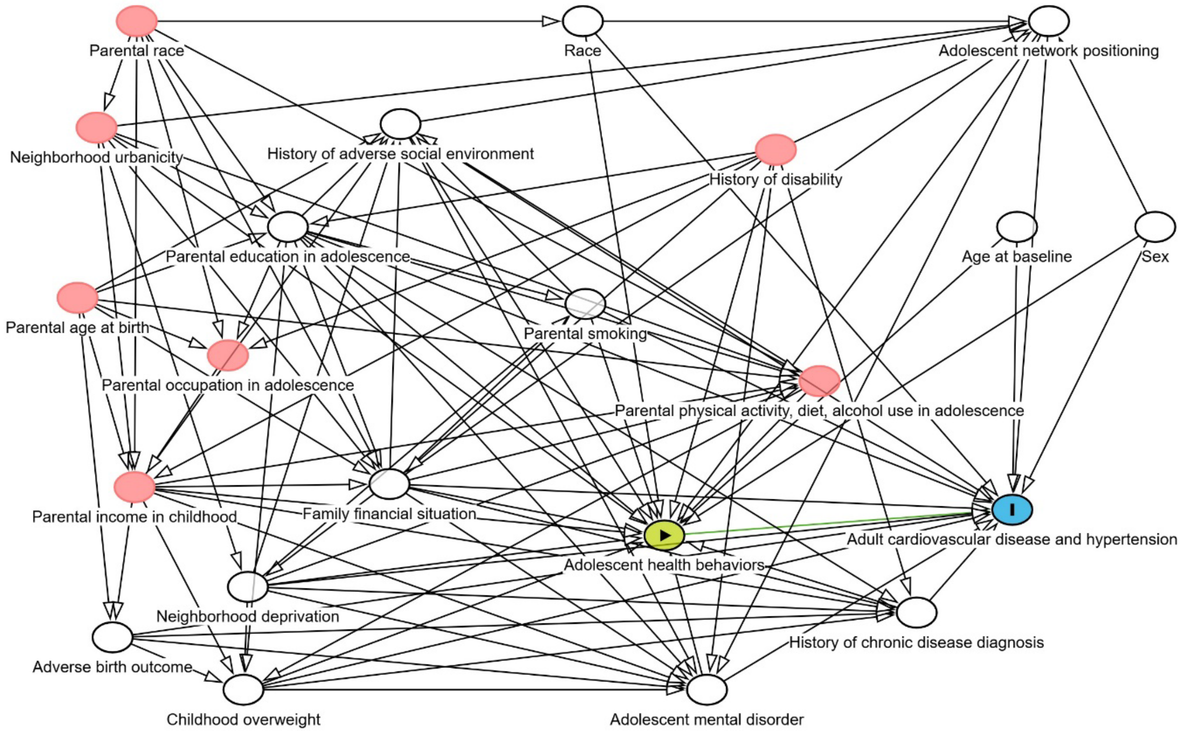


**Supplementary Fig. S1. Analytic DAG.** The exposure is marked with a triangle in a green circle and the outcome with an I in a blue circle. Variables in white nodes mark the minimal sufficient adjustment set. The graph was drawn with DAGitty (1) and can be found here:

https://dagitty.net/dags.html?id=fQ92WoKt

### 2. Add Health Questions Used for Health Behaviors

**Supplementary Table S1. Add Health survey variables used to define adolescent health behavior exposures.**

| Behavior | Wave | Variable name(s) | Survey question |
|---|---|---|---|
| Sports frequency | I in-home | H1DA5 | During the past week, how many times did you play an active sport, such as baseball, softball, basketball, soccer, swimming, or football? |
| Exercise frequency | I in-home | H1DA6 | During the past week, how many times did you do exercise, such as jogging, walking, karate, jumping rope, gymnastics or dancing? |
| Cigarette smoking | I in-home | H1TO1 | Have you ever tried cigarette smoking, even just 1 or 2 puffs? |
| | | H1TO2 | How old were you when you smoked a whole cigarette for the first time? If you have never smoked a whole cigarette, enter "0" |
| | | H1TO5 | During the past 30 days, on how many days did you smoke cigarettes? |
| | | H1TO7 | During the past 30 days, on the days you smoked, how many cigarettes did you smoke each day? |
| Alcohol use | I in-home | H1TO12 | Have you had a drink of beer, wine, or liquor—not just a sip or a taste of someone else's drink—more than 2 or 3 times in your life? |
| | | H1TO15 | During the past 12 months, on how many days did you drink alcohol? |
| Breakfast habits | I in-home | | What do you usually have for breakfast on a weekday morning? |
| | | H1GH23A | Milk |
| | | H1GH23B | Coffee or tea |
| | | H1GH23C | Cereal |
| | | H1GH23D | Fruit, juice |
| | | H1GH23E | Eggs |
| | | H1GH23F | Meat |
| | | H1GH23G | Snack foods |
| | | H1GH23H | Bread, toast or rolls |
| | | H1GH23I | Other items |
| | | H1GH23J | Nothing |

### 3. Standardized Mean Differences

We assessed the balance of confounders between the exposed and unexposed groups by comparing standardized mean differences (SMDs) before and after inverse probability weighting. The SMDs were calculated across 100 imputed datasets, and weights were applied to adjust for measured confounders, including age, sex, race, family financial difficulties, neighborhood deprivation, and other individual and familial factors. After applying the weights, we checked the average SMDs to ensure that balance was achieved for each behavior exposure. We conclude that a good balance was observed for all confounders (SMD <0.1).

**2A. Alcohol use**

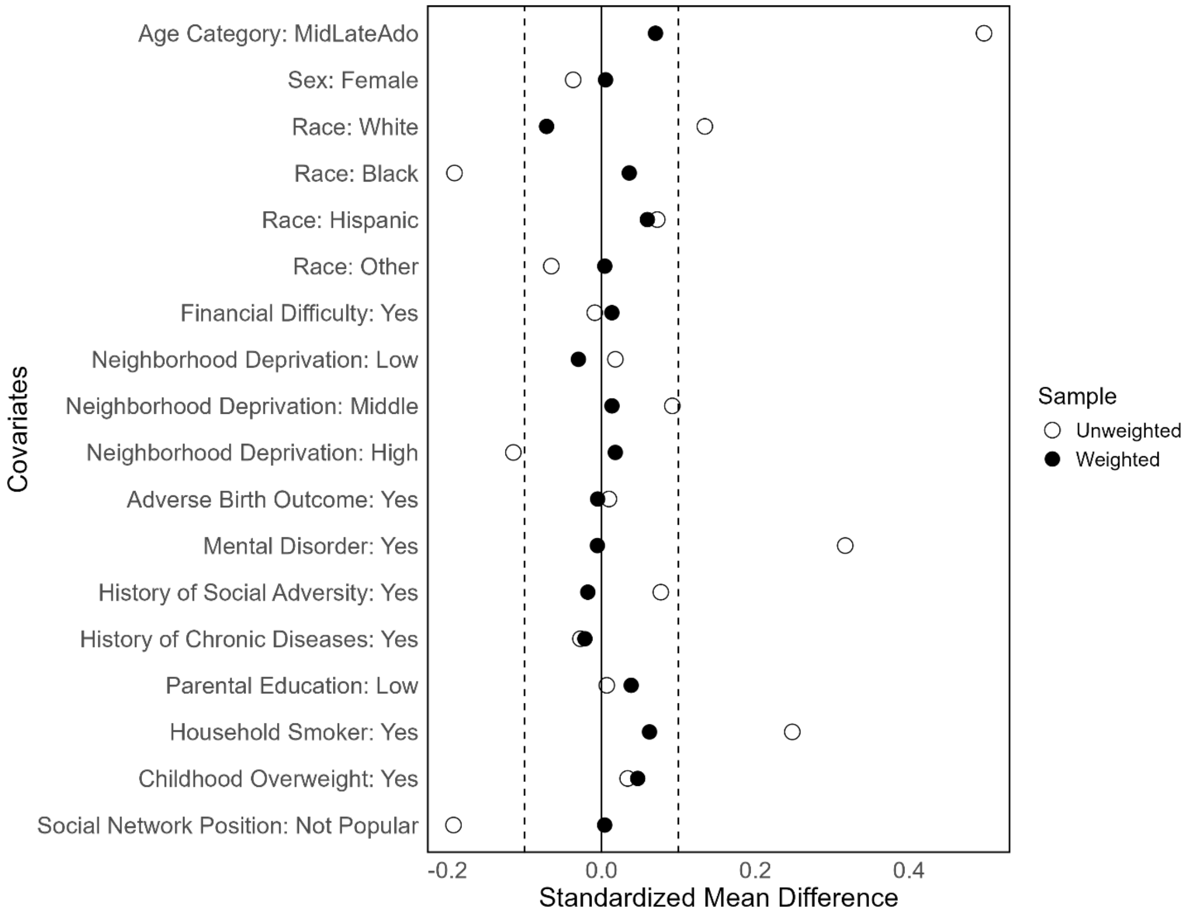

**2B. Breakfast habit**

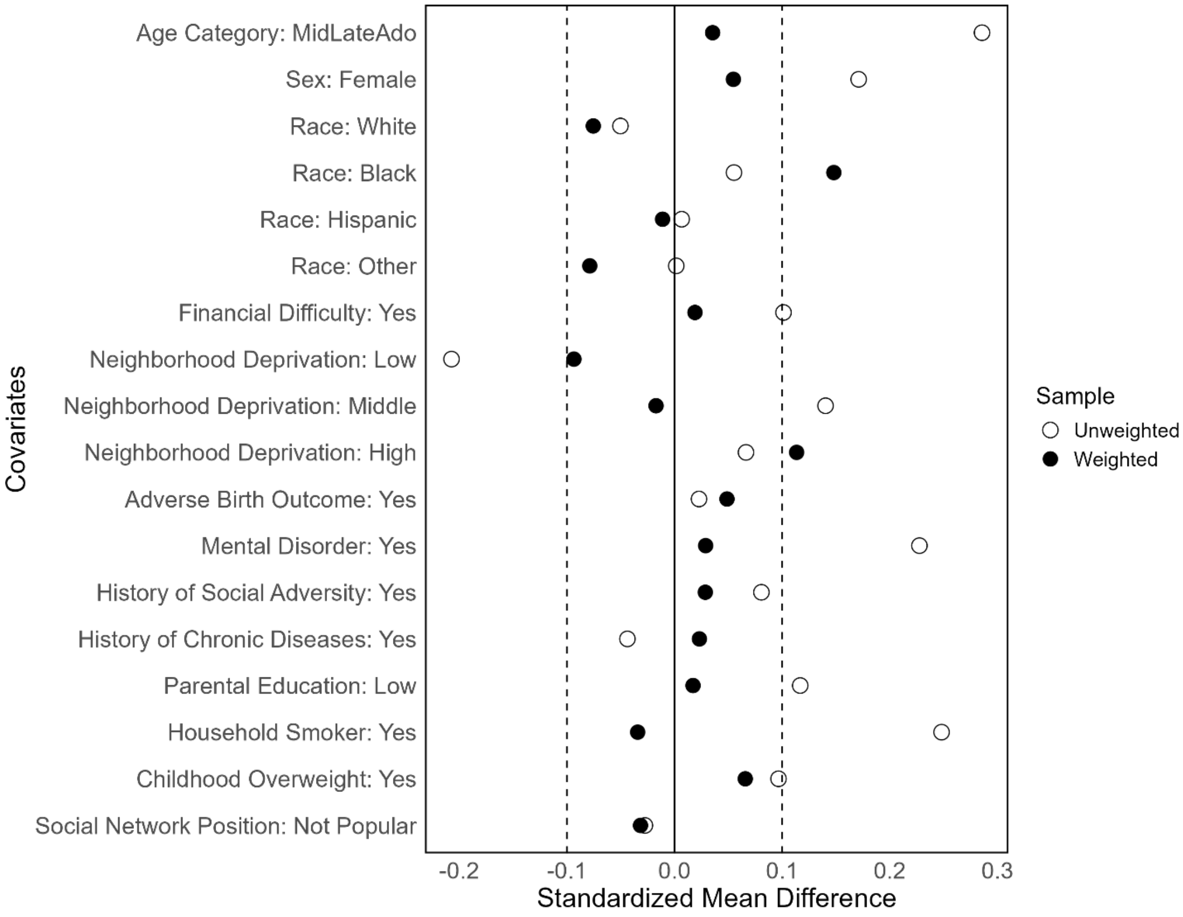

**2C. Exercise frequency**

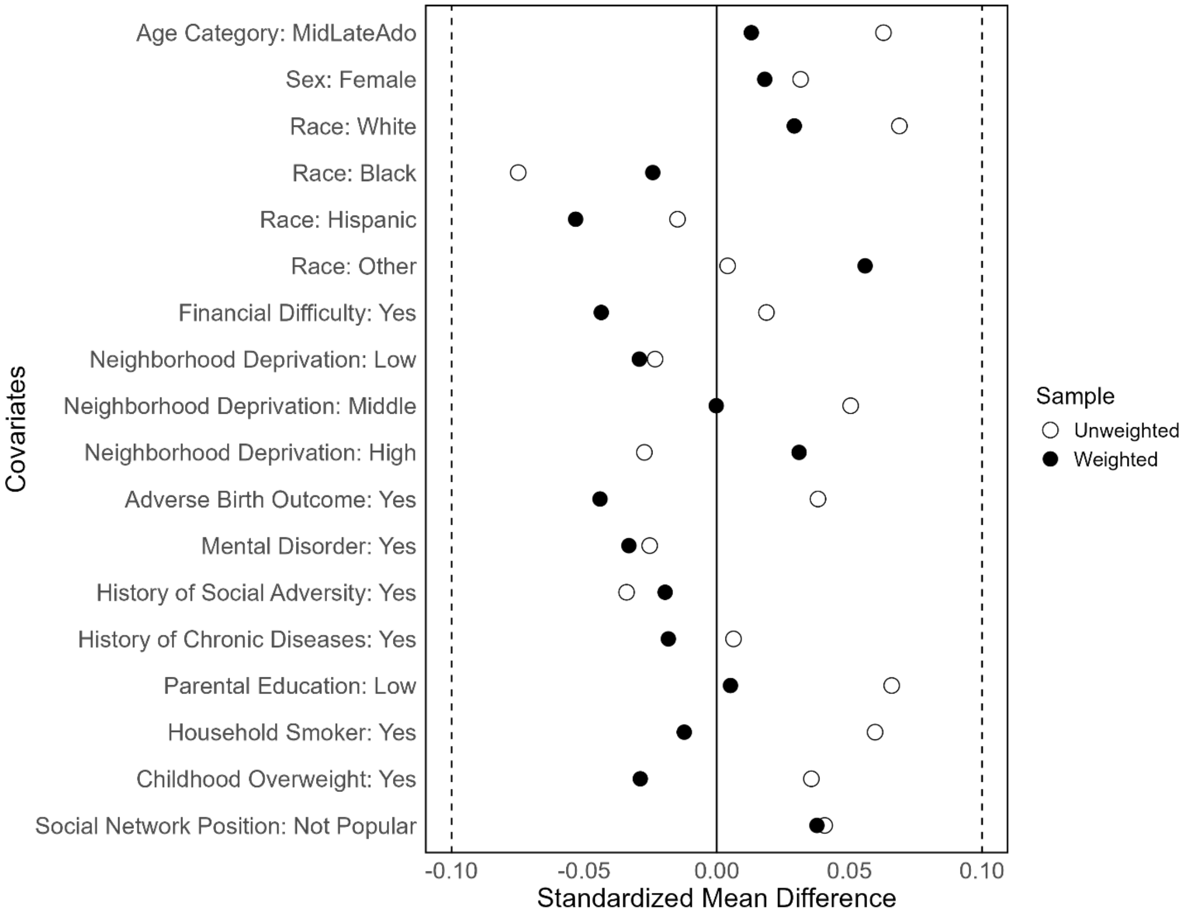

**2D. Smoking status**

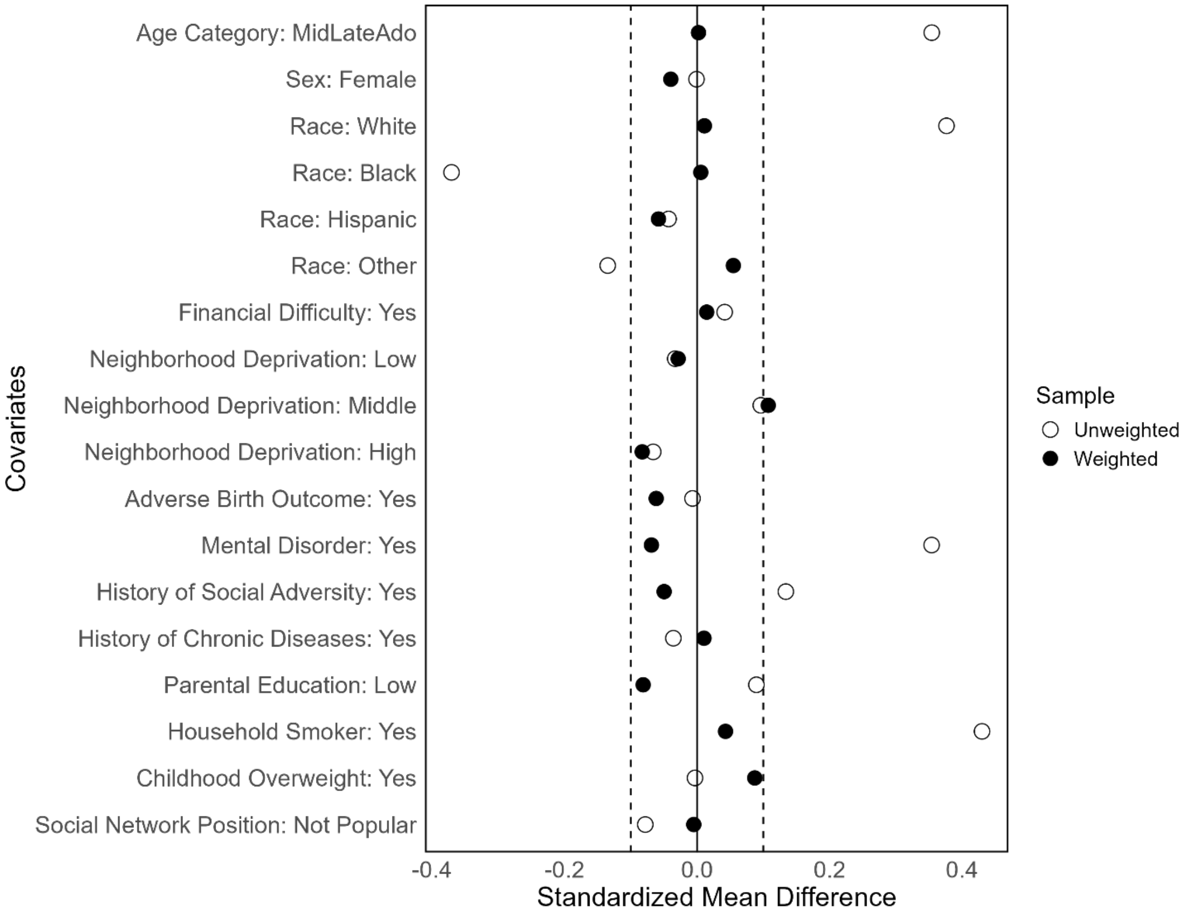

**2E. Sports frequency**

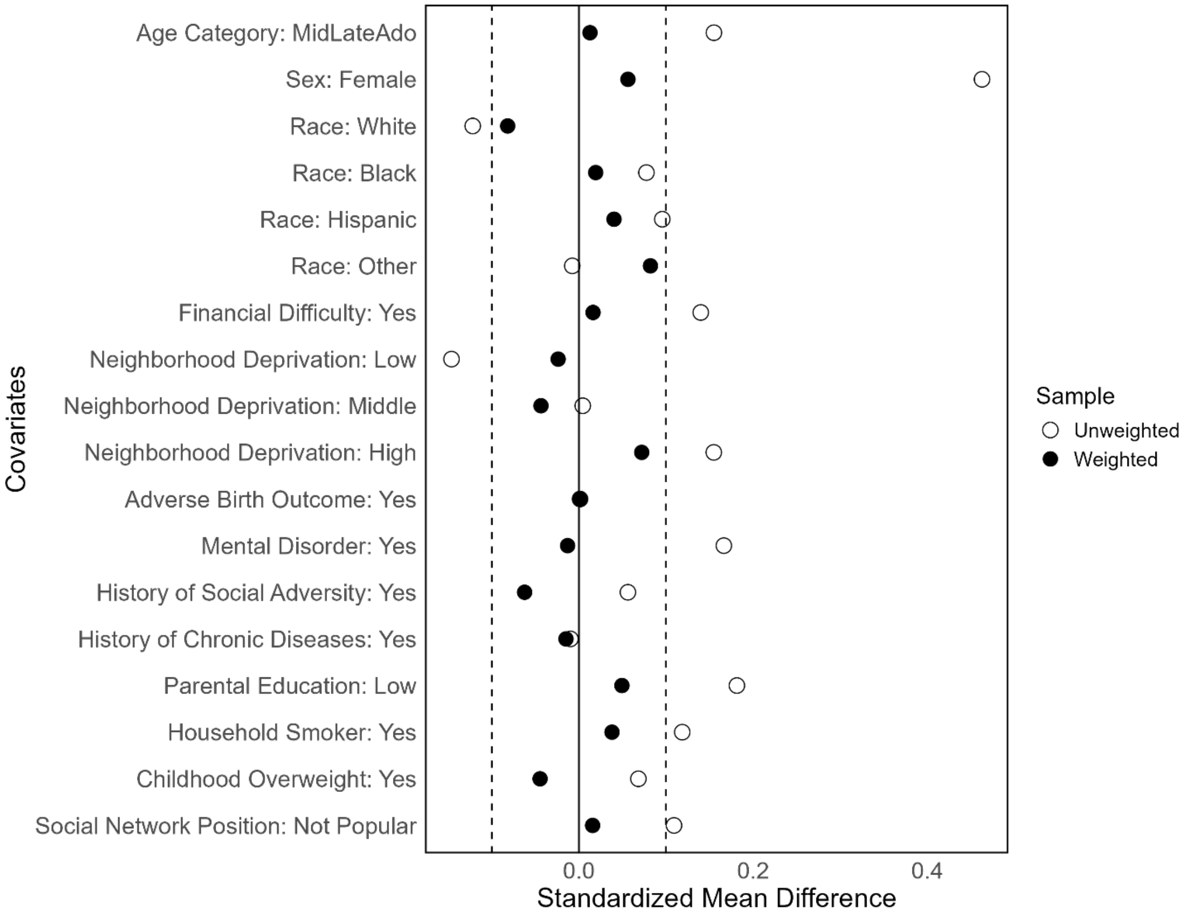

**2F. Alcohol use and concurrent smoking**

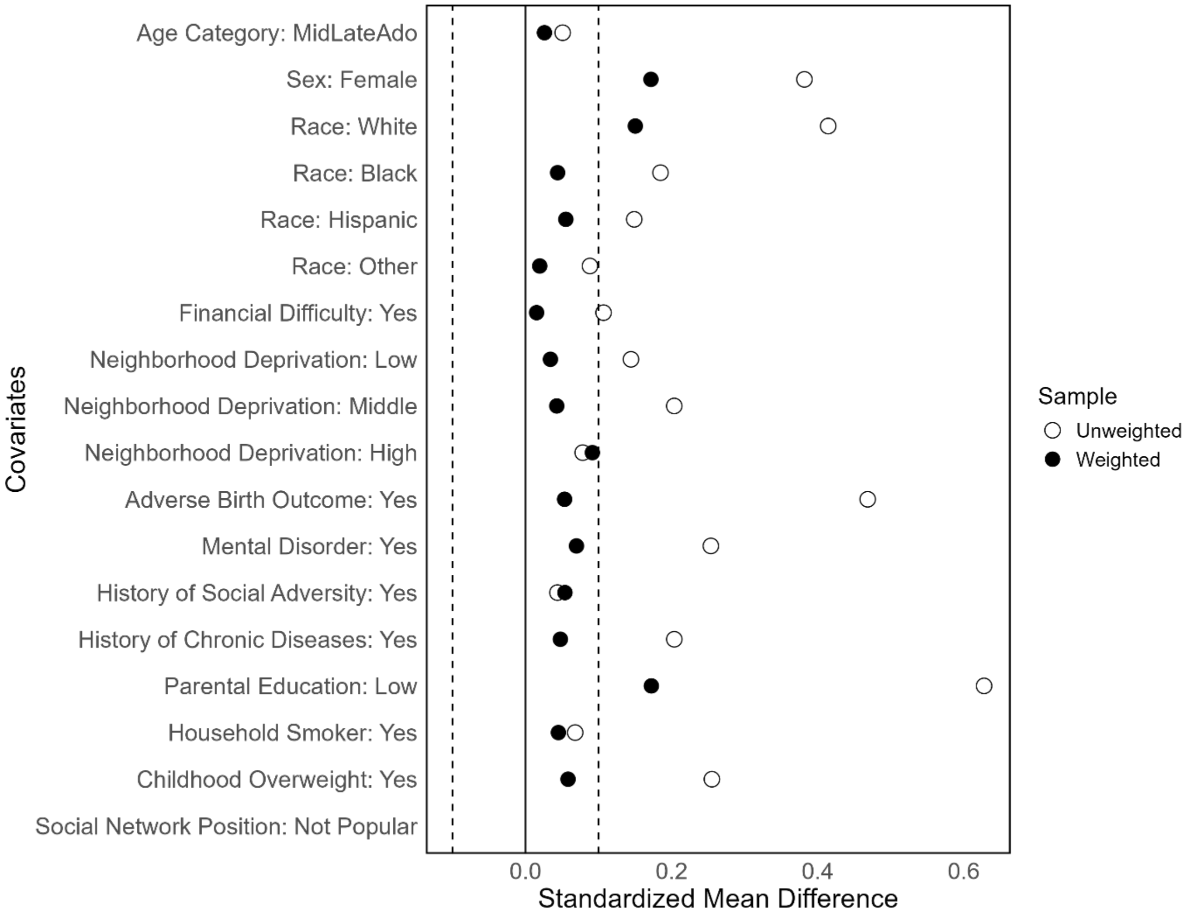


**Supplementary Fig. S2 A – F. Love plots of the covariance balance before and after inverse-probability of treatment weighting by treatment variables across the 100 imputed samples.** Dashed line corresponds to abs(SMD) = 0.1, values below are usually indicative of an achieved good balance.

### 4. Inverse Probability Weights and Imputation Models

We used inverse probability of the exposure weights in our causal analysis. Weights were calculated from a multinomial logistic regression model relating each exposure (dependent variable) to measured confounding factors (independent variables) specified as additive and linear without product terms. Final weights were truncated at 97.5% to minimize bias arising from few participants having large weights. To assess the weights distribution, we checked whether the means were roughly 1 and that none of the weights exceeded 10. We also assessed whether the weighted population corresponded to a population in which measured confounding factors are equally distributed across exposed and unexposed groups. This was done by comparing the standardized mean differences before and after weighting (see previous section).

We specified a separate marginal structural model for each outcome variable modelled as a function of the exposure, family financial difficulties, their product term, and the measured confounding factors as specified for the exposure model. As shown by Robertson et al., this guarantees our effect or differential effect estimates are double-robust (2). We assessed the goodness of fit of these marginal structural models by estimating the Homer-Lemeshow test and the natural course outcome risk. P values of the test ranged between 0.007 and 0.02 indicating a statistically significant goodness of fit. The natural course risk of the outcome (CVDs or hypertension) was 31.6% (95% CI: 30.1% – 33.1%) for alcohol use and similarly for the other exposures. These estimates align with the observed prevalence of the outcome (see Table 1), indicating a good prediction ability of the chosen outcome model specification (3).

To deal with missingness in variables that we hypothesized were missing at random (see Table 1 and Methods), we implemented multiple imputations by chained equations using the package MICE in R (4). For each bootstrap sample, we computed 100 imputed datasets and then pooled the cACEs applying Rubin’s rule (average). Predictors for the imputation were all complete covariates and three auxiliary variables that we identified in availability of cigarettes in the household reported by the adolescents or the interviewer (H1TO50 and H1IR23), whether the adolescent participant feels safe in their neighborhood (H1NB5), and whether the adolescent participant feels socially accepted (H1PF35).

### 5. Inequalities in Exposures and Outcomes

**Supplementary Table S2. Distribution of exposures and outcomes by effect modifier.**

Percentages (%) are weighted.

| | Family financial difficulty | | | |
|---|---|---|---|---|
| | Yes | | No | |
| | n | % | n | % |
| **Sports frequency** | | | | |
| High risk – infrequent sports | 484 | 66.2 | 2123 | 57.1 |
| Moderate risk – moderate frequency | 117 | 16.0 | 695 | 18.7 |
| Low risk – high frequency | 130 | 17.8 | 903 | 24.3 |
| **Exercise frequency** | | | | |
| High risk – infrequent exercise | 350 | 47.9 | 1789 | 48.1 |
| Moderate risk – moderate frequency | 181 | 24.8 | 876 | 23.5 |
| Low risk – high frequency | 200 | 27.4 | 1056 | 28.4 |
| **Cigarette smoking** | | | | |
| High risk – current smokers | 182 | 24.9 | 851 | 22.9 |
| Moderate risk – past smoker | 123 | 16.8 | 644 | 17.3 |
| Low risk – never smoker | 426 | 58.3 | 2226 | 59.8 |
| **Alcohol use** | | | | |
| High risk – moderate/frequent drinker | 203 | 27.8 | 1063 | 28.6 |
| Moderate risk – rare/past drinker | 205 | 28.0 | 955 | 25.7 |
| Low risk – never drinker | 323 | 44.2 | 1703 | 45.8 |
| **Breakfast habit** | | | | |
| High risk – breakfast skipping | 170 | 23.3 | 701 | 18.8 |
| Low risk – not breakfast skipping | 561 | 76.7 | 3020 | 81.2 |
| **CVDs or hypertension** | | | | |
| Yes | 265 | 36.6 | 1097 | 29.7 |
| No | 460 | 63.4 | 2592 | 70.3 |
| Missing | 6 | | 32 | |

### 6. Latent Class Analyses

We examined the co-occurrence of behaviors within an adolescent across the participants. Sports frequency was operationalized in four levels (no sports/low/moderate/high), exercise frequency into four levels (no exercise/low/moderate/high), smoking status into four levels (multiple cigarettes daily/one cigarette daily/past smoker/never smoker), alcohol use into five levels (frequent drinker/moderately drinker/rare drinker/past drinker/never drinker), and breakfast habit into three levels (usually skips breakfast/usually has cereal at breakfast/usually has some other food for breakfast).

Latent class analysis aims to establish a categorical grouping (or latent trait) that underlies patterns of responses across the different behaviors of sports, exercise, smoking, alcohol use and diet. We fitted models from 1 – 4 classes. The latent class analysis was carried out using the gsem package in Stata 18. We compared the model fit for each of the model results (Supplementary Table S4). The conditional probabilities of health behaviors for the two-class solution, three class solution and four class solution are graphed in Supplementary Figure S3.

**Supplementary Table S3. Model fit for 1 – 4 latent classes, n = 18,400.** The best values that guided the choice of the model are in bold.

| **Model** | **Log-likelihood** | **Resid. df** | **AIC** | **BIC** |
|---|---:|---:|---:|---:|
| Model 1 | -138994153.2 | 15 | 277988339 | 277988464 |
| Model 2 | **-**135311075 | 31 | **270622215** | **270622466** |
| Model 3 | -135566156 | 47 | 271132409 | 271132786 |
| Model 4 | -136206540 | 63 | 271413208 | 271413710 |

**3A. Two class solution**

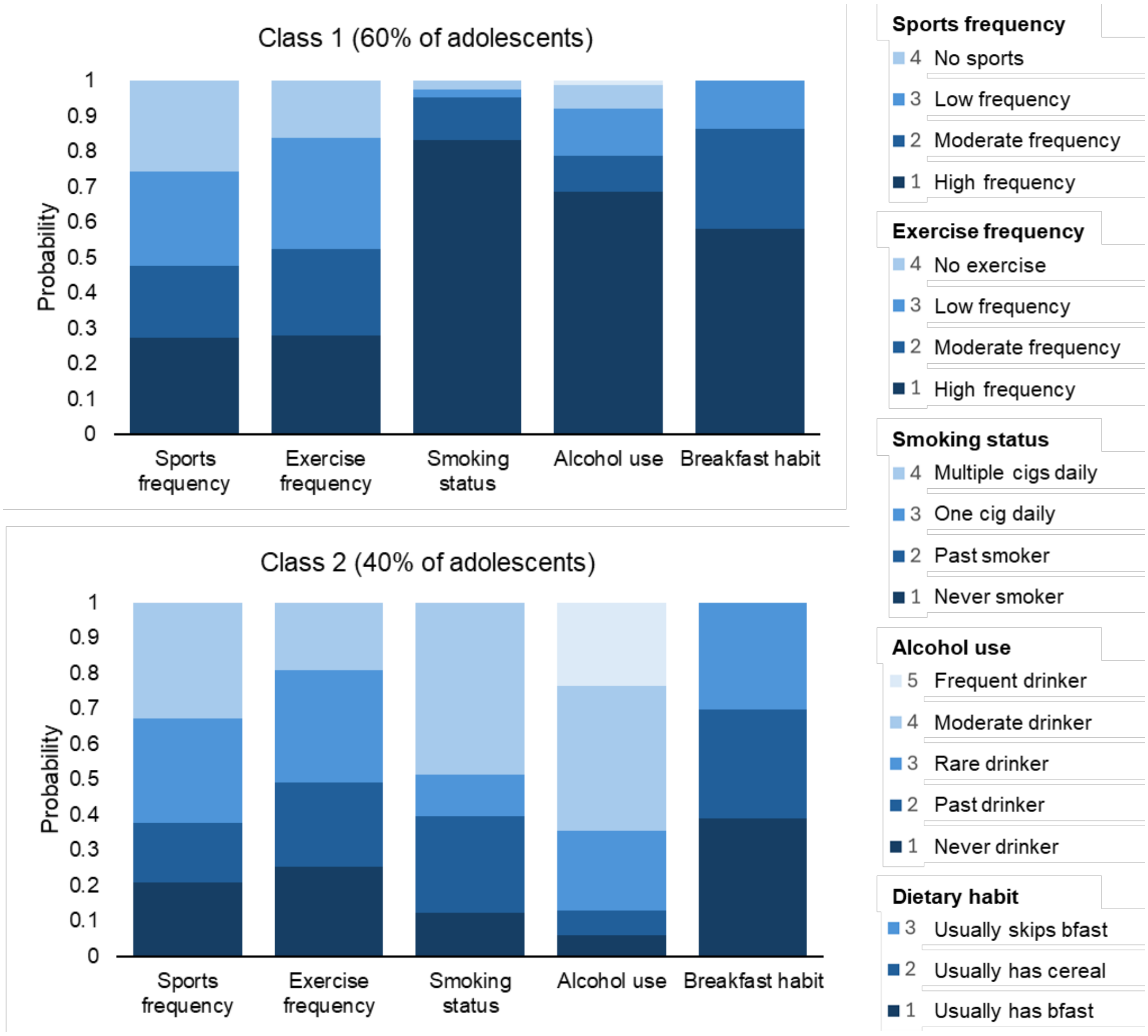

**3B. Three class solution:**

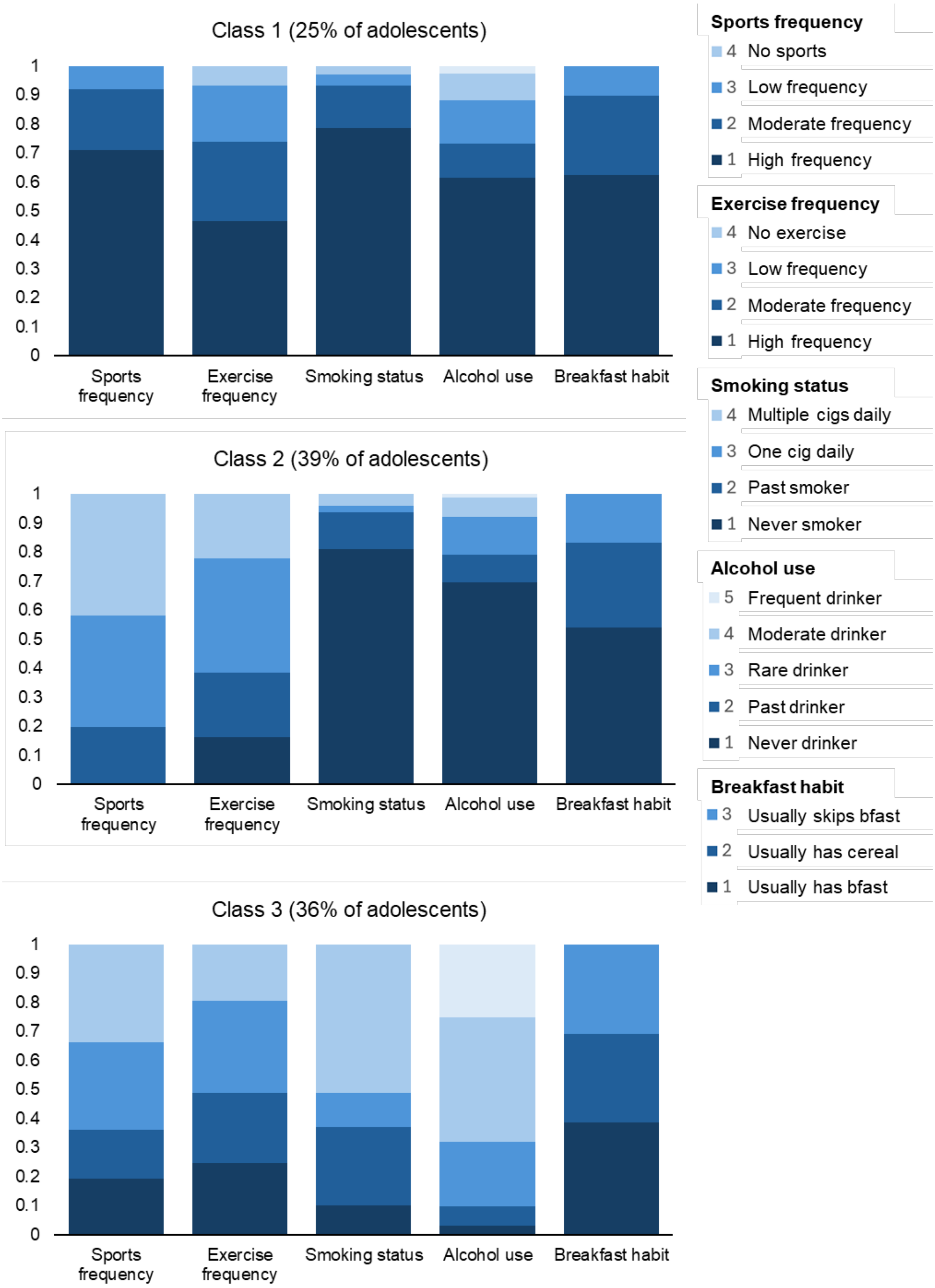

**3C. Four class solution:**

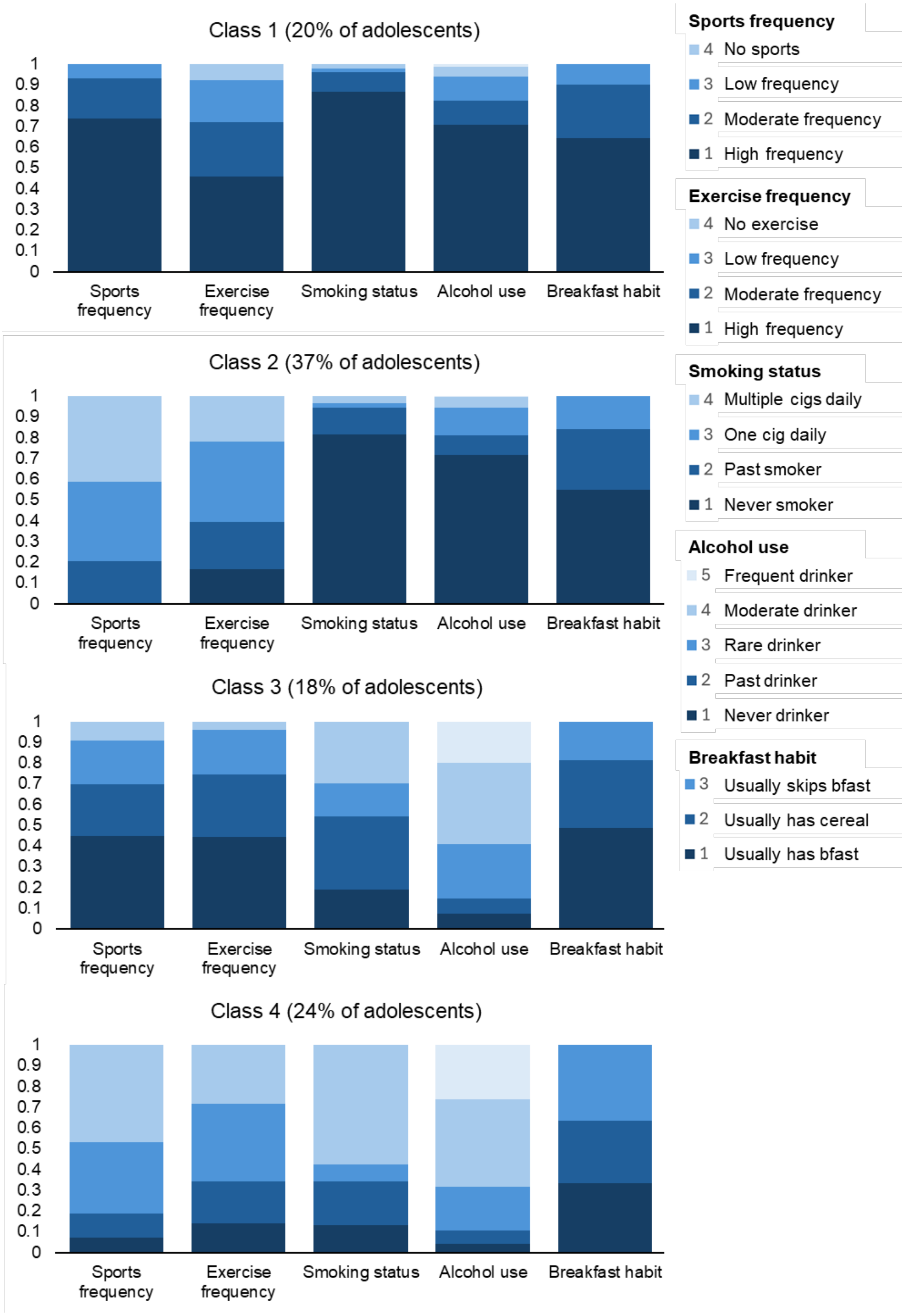


**Supplementary Figure S3. Conditional probabilities of health behaviors in a two class solution, a three class solution and a four class solution.**

### 7. Sensitivity Analyses

**Using equivalized parental income (top income quintile vs bottom income quintile)**
To assess the sensitivity of findings when using a different operationalization of the effect modifier, we ran the analysis when financial situation was measured through levels of equivalized parental gross income (bottom quintile vs top quintile) (n=4044).

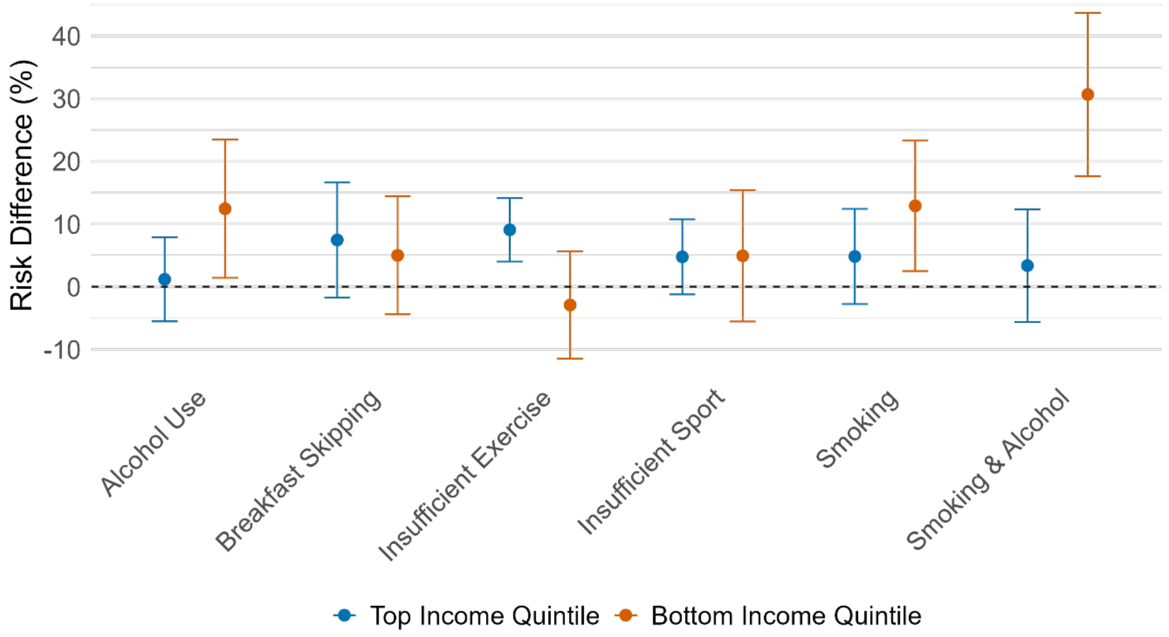


**Supplementary Fig. S4. Average causal effect of high risk health behaviors on cardiovascular conditions or hypertension among adolescents with higher parental income (top income quintile) and among adolescents with lower parental income (bottom income quintile).** Magnitude is measured via risk difference per 100 persons (% percentage).

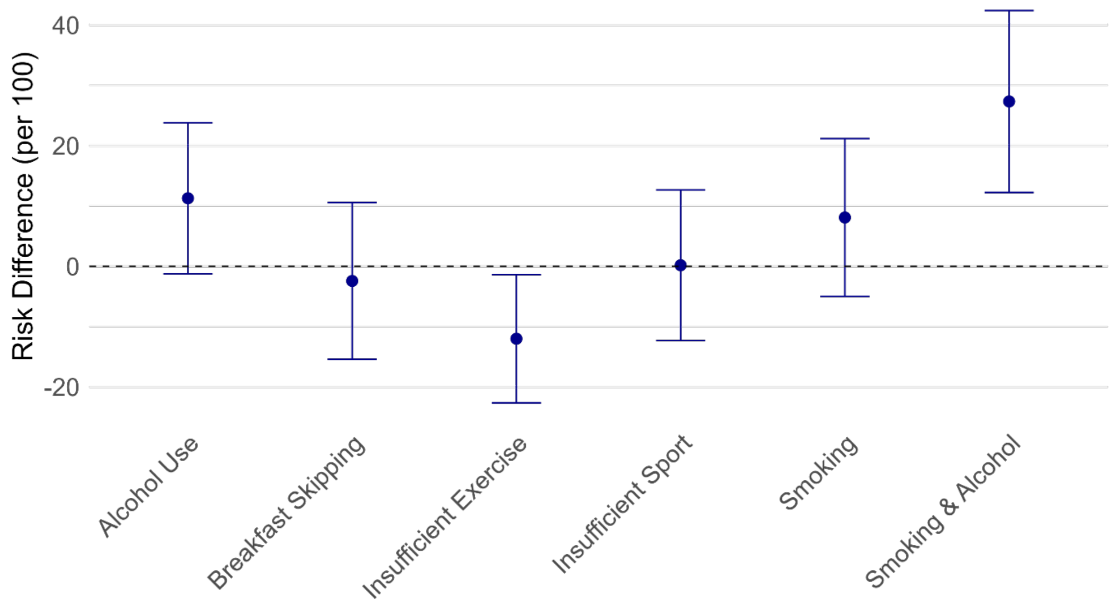


**Supplementary Fig. S5. Effect modification due to parental income.** The effect modification is the difference in average causal effect between those with high parental income (top income quintile) compared to those with low parental income (bottom income quintile).

**Negative control outcome**

To assess the sensitivity of our causal estimates to unmeasured confounding, we estimated differential effects on a negative control outcome that we identified in the self-reported voting participation in political elections. This outcome was derived from Wave V, where respondents were asked how often do they usually vote in local or statewide elections (variable H5SS8). For this analysis, we dichotomized the responses into two categories: "never" (those who responded that they never vote) and "usual" (those who reported voting sometimes, often or always).

**Supplementary Table S4**. Differential susceptibility to high risk health behaviors when the outcome is never/ever voting in political elections. Point estimate and 95% compatibility intervals are reported.

| High risk health behaviors – Exposure | Effect among adolescents from families without financial difficulty | Effect among adolescents from families with financial difficulty | Differential effect |
|---|---|---|---|
| Frequent/moderately frequent alcohol use | 3.8% (0.6% – 7.1%) | -0.8% (-8.2% – 6.7%) | -4.6% (-12.7% – 3.6%) |
| Breakfast skipping | -0.9% (-4.0% – 2.1%) | 8.2% (0.5% – 15.8%) | 9.1% (0.6% – 17.6%) |
| None/low frequency exercise | 1.6% (-1.3% – 4.4%) | 3.1% (-4.1% – 10.3%) | 1.6% (-6.1% – 9.2%) |
| None/low frequency sport | 0.8% (-2.5% – 4.1%) | -3.0% (-11.3% – 5.2%) | -3.8% (-13.1% – 5.4%) |
| Current smoking | 4.8% (1.6% – 8.0%) | 5.5% (-2.8% – 13.8%) | 0.8% (-8.3% – 9.6%) |
| Smoking & alcohol use | 3.8% (-0.6% – 8.1%) | 4.3% (-5.3% – 13.9%) | 0.5% (-10.3% – 11.4%) |

### 8. Main Results Tables

**Supplementary Table S5.** Point estimate and 95% compatibility intervals for the cACE and the effect modification of high risk behaviors on cardiovascular diseases or hypertension, conditional on socioeconomic family financial situation as displayed in Figure 3 and 4.

| Outcome | Effect among adolescents from families without financial difficulty | Effect among adolescents from families with financial difficulty | Effect modification |
|---|---|---|---|
| Frequent/Moderately frequent alcohol use | 1.1 (-2.0 – 4.3) | 10.8 (2.3 – 19.2) | 9.7 (0.8 – 18.5) |
| Breakfast skipping | 4.7 (0.7 – 8.7) | 10.1 (1.6 – 18.7) | 5.4 (-3.7 – 14.6) |
| None/low frequency exercise | -1.7 (-4.7 – 1.2) | 0.8 (-6.7 – 8.3) | 2.6 (-5.5 – 10.7) |
| None/low frequency sport | 2.3 (-1.1 – 5.8) | 2.2 (-6.9 – 11.2) | -0.2 (-9.8 – 9.5) |
| Current smoking | -0.2 (-3.6 – 3.2) | 1.8 (-6.7 – 10.3) | 2.0 (-7.2 – 11.2) |
| Smoking & alcohol use | 0.4 (-4.1 – 4.8) | 11.4 (0.9 – 22.0) | 11.1 (0.1 – 22.1) |

**Supplementary Table S6.** Point estimate and 95% compatibility intervals for the cACE and the effect modification of moderate risk behaviors on cardiovascular diseases or hypertension, conditional on socioeconomic family financial situation. Breakfast skipping is omitted as this exposure was dichotomized into high risk and low risk.

| Outcome | Effect among adolescents from families without financial difficulty | Effect among adolescents from families with financial difficulty | Effect modification |
|---|---|---|---|
| Rare/Past alcohol use | -0.9 (-4.4 – 2.6) | 4.3 (-3.6 – 12.2) | 5.2 (-3.4 – 13.7) |
| Moderately frequent exercise | 0.5 (-3.8 – 4.9) | -2.4 (-11.1 – 6.2) | -3.0 (-13.0 – 7.0) |
| Moderately frequent sport | 2.3 (-2.5 – 7.1) | 11.8 (-0.6 – 24.3) | 9.5 (-2.9 – 21.9) |
| Past smoking | 3.2 (-1.3 – 7.7) | 6.8 (-3.9 – 17.4) | 3.5 (-8.4 – 15.5) |

**Supplementary Table S7.** Point estimate and 95% compatibility intervals for the cACE and the effect modification of high risk behaviors on hypertension only, conditional on socioeconomic family financial situation.

| **Outcome** | **Effect among adolescents from families without financial difficulty** | **Effect among adolescents from families with financial difficulty** | **Effect modification** |
|---|---|---|---|
| Frequent/moderately frequent alcohol use | 1.2 (-2.1 – 4.4) | 8.5 (0.3 – 16.7) | 7.4 (-1.6 – 16.3) |
| Breakfast skipping | 5.2 (1.1 – 9.3) | 10.7 (2.2 – 19.2) | 5.5 (-3.7 – 14.7) |
| None/low frequency exercise | -0.6 (-3.6 – 2.4) | 1.9 (-5.3 – 9.1) | 2.5 (-5.5 – 10.5) |
| None/low frequency sport | 2.3 (-1.4 – 6.0) | 2.2 (-7.0 – 11.5) | -0.1 (-9.9 – 9.7) |
| Current smoking | 0.1 (-3.2 – 3.3) | 2.6 (-5.5 – 10.6) | 2.5 (-6.3 – 11.3) |
| Smoking & alcohol use | 1.0 (-3.5 – 5.4) | 11.4 (1.0 – 21.9) | 10.5 (-0.7 – 21.7) |
